# The mystery of COVID-19 reinfections: A global systematic review and meta-analysis of 577 cases

**DOI:** 10.1101/2021.07.22.21260972

**Authors:** Rubaid Azhar Dhillon, Mohammad Aadil Qamar, Omar Irfan, Jaleed Ahmed Gilani, Usama Waqar, Mir Ibrahim Sajid, Syed Faisal Mahmood

## Abstract

**Background:** As the COVID-19 pandemic rages on, reports on disparities in vaccine roll out alongside reinfection and reactivation from previously recovered cases have been emerging. With newer waves and variants of COVID-19, we conducted a systematic review to assess the determinants and disease spectrum of COVID-19 reinfection.

**Methods:** A comprehensive search covering relevant databases was conducted for observational studies reporting Polymerase Chain Reaction (PCR) confirmed infection and reinfection cases. Quality assessment tool developed by the National Institute of Health (NIH) for assessment of case series was used. Meta-analyses were performed using RevMan 5.3 for pooled proportions of findings in first infection and reinfection with 95% confidence interval (CI).

**Results:** Eighty-one studies reporting 577 cases were included from 22 countries. The mean age of patients was 46.2±18.9 years with males accounting for 45.8% of the study population while 179 (31.0%) cases of comorbidities were reported. The average time duration between first infection and reinfection was 63.6±48.9 days. During first infection and reinfection, fever was the most common symptom (41.4% and 36.4%, respectively) whilst anti-viral therapy was the most common treatment regimen administered (44.5% and 43.0%, respectively). Overall, comparable odds of symptomatic presentation and management were reported in the two infections. However, a higher Intensive Care Unit (ICU) admission rate was observed in reinfection compared to first infection (10 vs 3). Ten deaths were reported with 565 patients fully recovering. Respiratory failure was the most common cause of death (7/10 deaths). Seventy-two studies were determined to be of good quality whilst nine studies were of fair quality.

**Conclusion:** As the first global-scale systematic review of its kind, our findings support immunization practices given increased ICU admissions and mortality in reinfections. Our cohort serves as a guide for clinicians and authorities for devising an optimal strategy for controlling the pandemic.

## Introduction

The coronavirus, through its rapid spread and emerging variants, started in Wuhan amid December 2019. It was declared a global pandemic in March 2020 and persists as a public healthcare emergency. Over this period of more than a year, till date, the virus has infected more than 165 million individuals globally, and has resulted in more than 3.4 million deaths [1].

Currently, there are seven types of corona viruses known to infect humans: 4 are seasonal and cause limited upper respiratory tract infections, but 3 of them, namely SARS coronavirus (SARS-CoV-1), Middle East respiratory syndrome (MERS), and SARS-CoV-2 cause severe disease [2]. SARS COV-2, the cause of the current pandemic, causes infection by binding to the angiotensin converting enzyme 2 receptor using a receptor binding domain in its spike protein for cell entry and ultimately, resulting in a respiratory syndrome [2–4]. The currently available vaccines target the spike protein[2]. However, mutations in the spike protein have been implicated in the reduction of small, but significant, efficacy of vaccines [5, 6] highlighting the scale of the challenge COVID-19 places to the world.

Even though the world is heralding the development of new vaccines as a potential way forward for a healthy, more “COVID-free” time, the concern of reinfection, recurrence, and mutant variants looms. The Center for Disease Control and Prevention (CDC), in their 27 ^th^ October 2020 update[7] raised concerns over reactivation of the disease and requested the public to maintain infection control measures, such as wearing a mask in public, maintaining a six feet distance, regular hand washing, and avoiding crowded spaces.

Since Tillett et al.,[8] described the first confirmed case of COVID-19 reinfection from the USA, many authors have described their patient experiences of viral recurrence. The reason for this recurrence and its potential public health implication is a question that warrants explanation. Iwasaki et al.,[9] reason that perhaps a scant antibody response following the first infection could be the cause of a relapse. They emphasize on ascertaining a degree of specificity of the antibody (anti-nucleocapsid vs anti-spike antibody) at the time of reinfection, as well as determine the immune correlations of protection.

The public concern of whether vaccines could be a potential cure for the viral outbreak remains to be explained, with the obvious apprehension of whether a separate vaccine would be required for every variant of the virus. Till date, they have been four variants with significant mutations in the spike protein that have gained wide spread surveillance: B.1.1.7 (VOC 202012/01 or 20B/501Y.V1) which originated in the UK, B.1.351 (20H/501Y.V2) which originated from the Republic of South Africa, and P.1 (B.1.1.28.1) which was reported in travelers coming from Brazil [10]. A fourth double mutant variant of concern, labeled B.1.617 coming out from India, has been reported in March 2021 and is being investigated, considered to be a cause of the massive rise in infections in India currently experiencing its second wave [11].

A few questions remain though. Firstly, would immunity conferred by the first infection protect individuals from a serious disease process in the reinfection phase? And secondly, does reinfection imply that individuals who are already vaccinated experience a more severe COVID-19 infection? The objective of this review, in addition to providing a comprehensive evidence on COVID-19 reinfections including both pediatric and adult cases, is a unique comparison of first infection and the reinfection disease spectrum, management, and outcomes.

## Methods

The protocol of the review is registered with PROSPERO CRD42021239816. The review is reported in accordance with the Preferred Reporting Items for Systematic Reviews and Meta-Analyses (PRISMA).

### Search methods

An exhaustive literature review was conducted on major databases: PubMed, WHO COVID-19 Database, Embase, China National Knowledge Infrastructure (CNKI) Database, Google Scholar, manual searches of leading medical journals, and a pre-print server, medRxiv, covering the timeline of January 1^st^, 2020, to March 16^th^, 2021. The following keywords were used to conduct the search: COVID-19 and derivatives, reinfection, relapse, reactivation, as shown in S1 Table. Complementary searches were conducted in the John Hopkins Health Resource, Chinese and US CDC Library. No language restrictions were applied. Key reference lists were additionally screened for more studies.

### Selection strategy

Observational studies (cohorts, case series, and case reports) reporting laboratory-confirmed COVID-19 (RT-PCR) first infection and reinfection were considered for inclusion. Review articles, commentaries, and letters not presenting any original data were excluded. Covidence software (2016 edition) was used for screening by two reviewers independently and in duplicate. Any discrepancies in selection were resolved by a third independent author.

### Data extraction and analysis

The shortlisted articles were then extracted independently and in duplicate, on a structured data form by two reviewers. The information extracted was as follows: Author names, date of publication, country, setting of study, type of study, number of patients and age group, patient information (age, gender and comorbidities), clinical evaluation (presenting symptoms in both infections, travel/exposure history and infected family members), diagnostic tests (nasopharyngeal swabs, antibody tests and timelines for initial infection and reinfection), radiographic findings, therapeutic regimen (medications, isolation, and plasma therapy), outcomes for both infections (hospitalization, ICU admission, complications, discharge, and death) and antibody status after both infections. Disaggregated data by age groups (children and adults) was extracted where available.

Categorical data was summarized as counts and proportions. The pooled proportions of reported findings were calculated using Review Manager 5.3’s random-effects model. I^2^ was calculated to examine statistical heterogeneity. The clinical features and outcomes were compared accordingly between first infection and reinfection using pooled proportions and their 95% CIs, supplemented by an odds ratio (OR).

### Quality assessment

Quality assessment of included studies was conducted using the tool developed by the National Institute of Health (NIH) for assessment of case series. Two authors independently scored the quality of case series out of 8, based on the clarity of study objectives, case definition, consecutive subject recruitment, comparability of subjects, definition and measurement of outcomes, length of follow-up, statistical methods, and results. A case series scoring 6-8 was considered good quality, 4-5 was considered fair quality and <4 was considered poor quality. Consecutive subject recruitment, comparability of subjects and statistical methods were excluded from the scoring criteria for case reports, which were scored out of 5. Case reports scoring 5 were considered good quality, scoring 3-4 were considered fair quality, and scoring <3 were considered poor quality.

## Results

The comprehensive literature search yielded a total of 980 studies after removal of duplicates. After title and abstract screening, 87 studies were found to be eligible for a full-text assessment, of which, 81 were found to be eligible according to our inclusion criteria[8, 12–91]. The characteristics of the 81 included studies is shown in S2 Table. Six studies were excluded as they presented overlapping or missing data. An overview of the detailed systematic study selection process is presented in the PRISMA flow diagram (Fig 1).

**Fig 1.**
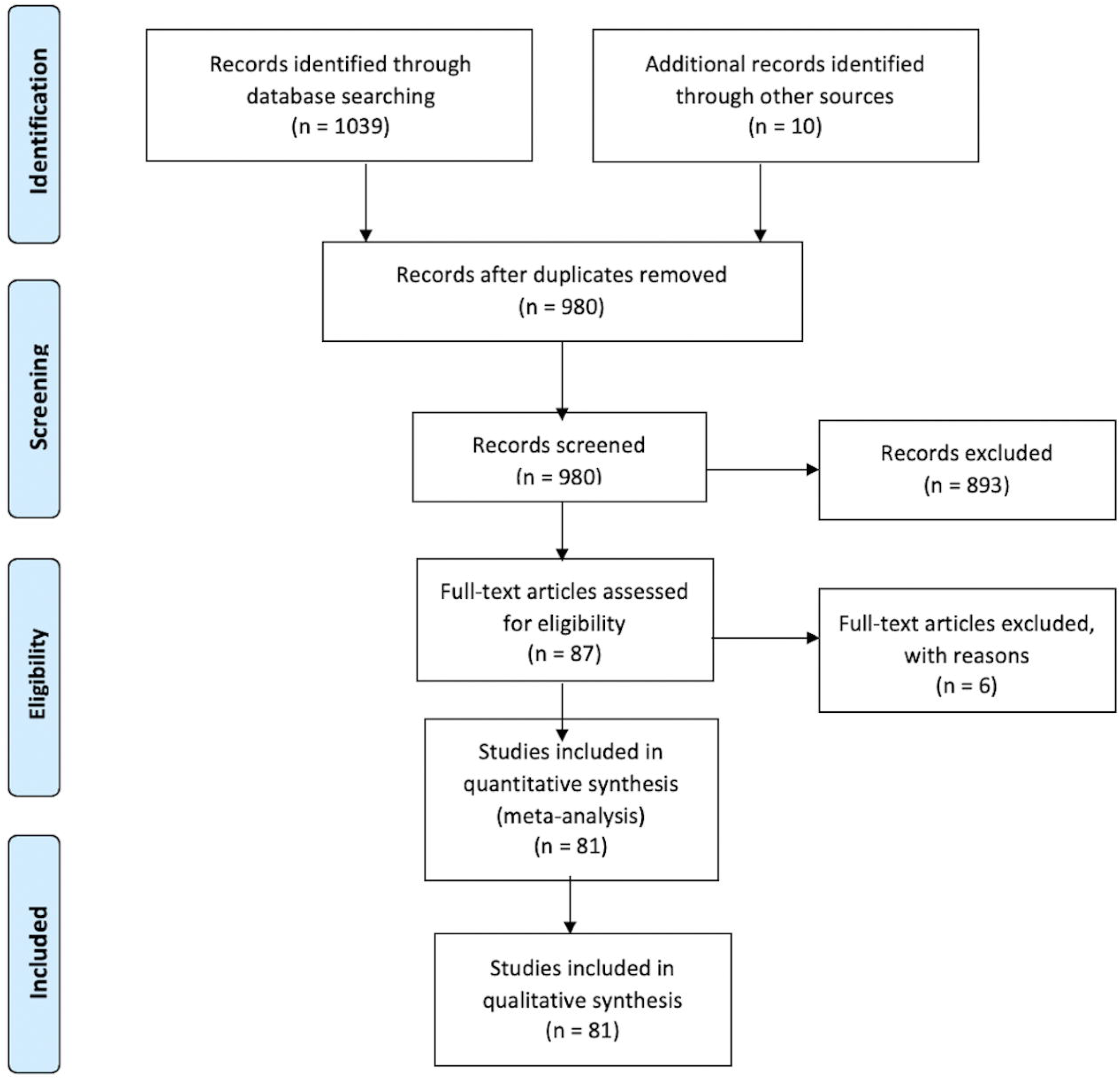
PRISMA Flow diagram of study selection process.

Through consultation with a professional librarian, a search was conducted in leading medical journals and through duplication and screening processes, studies were selected.

In terms of study types, 45 studies (55.6%) were case reports, and 36 studies (44.4%) were case series. Studies were reported from 22 countries. Forty-eight out of 81 studies were reported from low-and-middle-income countries (LMICs). More than one-third of the studies were reported from China (28/81, 34.6%), almost one-sixth were from the USA (11/81, 13.6%), accounting together for about half of the studies (39/81, 48.1%) included in our review. The global distribution of studies is shown in Table 1 and S1 Fig.

**Table 1.** Distribution of studies (N=81) and reinfection cases (N=577) according to country of origin included in the review.

| Country | Number of studies | Number of reinfection cases |
| --- | --- | --- |
| Belgium | 2 | 2 |
| Brazil | 7 | 45 |
| China | 28 | 423 |
| Colombia | 1 | 1 |
| Ecuador | 1 | 1 |
| France | 5 | 19 |
| Hong Kong | 3 | 3 |
| India | 4 | 8 |
| Iraq | 1 | 26 |
| Israel | 1 | 1 |
| Italy | 4 | 4 |
| Iran | 1 | 9 |
| Netherlands | 1 | 1 |
| Pakistan | 1 | 1 |
| Peru | 1 | 1 |
| Portugal | 1 | 1 |
| Qatar | 1 | 1 |
| UK | 3 | 8 |
| USA | 11 | 14 |
| Turkey | 2 | 3 |
| Switzerland | 1 | 1 |
| Korea | 1 | 4 |
| **Total** | **81** | **577** |

### Demographics and Epidemiology

A total of 577 cases with a mean age of 46.2±18.9 years (range 3.0 – 91.0 years) were included in the study. The gender of study cases was noted to be 45.8% males and 53.7% females. Of the 577 cases, approximately one-third of the cases (n=179, 31.0%) were reported to have at least one comorbidity. Reports of having a positive contact history with a close contact or a family member with COVID-19 were found in 87 (15.1%) cases (S2 Table and Table 2). Across 76 studies, the average reported time duration between first infection and reinfection was 63.6 ± 48.9 days (range 11.0 – 210.0 days).

**Table 2.** Demographics of Patients.

| Variable | Mean Proportion (%) |
| --- | --- |
| **Age (Years)** | 46.2 ± 18.9 years (range 3.0 – 91.0 years) |
| **Gender** |  |
| Male | 264 (45.8) |
| Female | 310 (53.7) |
| Not specified | 3(0.5) |
| **Contact History Positive** | 87 (15.1) |
| **Underlying Comorbidities** | 179 (31.0) |
| **Average time duration between first infection and reinfection** | 63.6 ± 48.9 days (range 11.0 – 210.0 days) |

### Clinical features during first infection and reinfection

Around three-fourth of our cases were categorized as being mildly symptomatic with only 10 cases being classified as severe to critical by their respective studies for disease severity in the first infection. The total number of asymptomatic cases during the first infection was 53 (9.2%) with an increase to 184 (31.9%) cases noted during reinfection (S2 Table). Tian M et al.,[72] reported the highest number of asymptomatic cases (20/577, 3.5%) during the first infection whereas An J et al.,[17] with 27 cases (4.7%) reported the highest number of asymptomatic cases during reinfection. The presence of antibodies was also reported for the total 577 cases, wherein 323 (56.0%) and 364 (63.0%) cases were detected to be positive during first infection and reinfection, respectively.

The most common presenting symptoms amongst patients in the first infection were fever (n=239, 41.4%) and cough (n=201, 34.8%), which then accounted for 36.4% (n=210) and 34.8% (n=201) of cases, respectively, during reinfection. Myalgia was reported in 80 (13.9%) cases during first infection which increased to 88 (15.3%) cases during reinfection. The frequencies and odds ratio of other reported signs and symptoms are listed in Fig 2.

**Fig 2.**
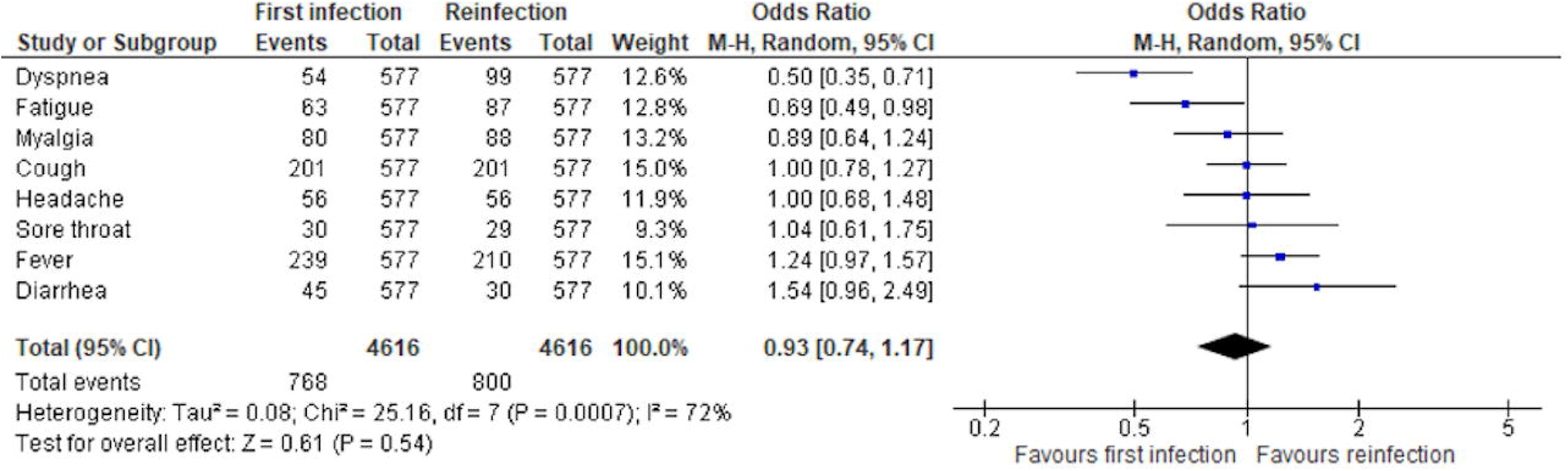
Clinical features of First Infection and Reinfection (N=577)

Regarding the radiological imaging during first infection of the 577 cases, almost one-quarter (146/577, 25.3%) had not reported, or did not have, any kind of chest imaging done. Out of those who reported, only 25 (4.3%) cases had a normal finding whereas 295 (51.1%) cases reported an abnormality with Chen et al., [27] reporting the highest occurrence of radiological abnormalities (70 out of 81 cases, 86.4%).

### Management of first infection and reinfection

The most administered treatment, as reported by the study results, was antiviral therapy accounting for 44.5% (n=257) and 43.0% (n=248) of cases during first infection and reinfection, respectively. Administration of antibiotics was lower at 14.4% (n=83) and 13.2% (n=76) for first infection and reinfection respectively (Fig 3).

**Fig 3.**
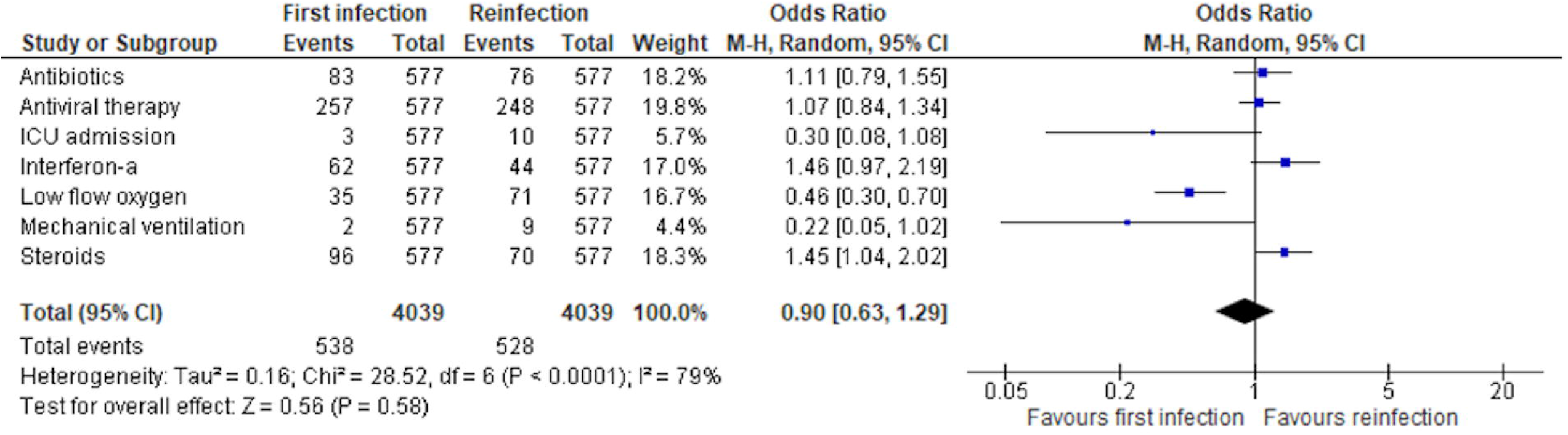
Management of First Infection and Reinfection (N=577)

In the study by He et al. [42], 60% of patients were administered steroids during both infections, which was reported to be the highest use of steroids amongst studies reported to date. However, in our data, the overall use of steroids stood at 16.6% (n=96) and 12.1% (n=70) during the respective infections. Traditional Chinese medicine and interferon administration were reported at 13.9% (n=80) and 10.7% (n=62) during the first infection and 26.7% (n=154) and 7.6% (n=44) during reinfections, respectively.

Lastly, our results show that the use of low flow oxygen stood at 6.1% (n=35) of cases during the first infection which then doubled to 12.3% (n=71) of cases during reinfection (Fig 3). Out of the total studies, reports of ICU admission and mechanical ventilation was relatively low being 0.5% (n=3) and 0.3% (n=2) during first infection, respectively, compared to 1.7% (n=10) and 1.6% (n=9) during reinfection, respectively. This was contradictory to a higher number of asymptomatic cases observed in the reinfection phase, implicating for a possible need of an Individual Patient Data (IPD) analysis in future studies.

Comparable odds of symptomatic presentation (OR:0.93, 95%CI: 0.74-1.17) and management (OR:0.90, 95%CI: 0.63-1.29) were observed in the first infection compared to reinfection when meta-analyzed, as shown in Fig 2 and Fig 3. Although a higher event of management was observed in the first infection, due to the individual weight of the studies, the overall OR favored first infection and was less than 1.

### Outcomes of first infection and reinfection

Complete recovery rate after reinfection stood at 97.9% (565 cases) with a total of 10 (1.8%) deaths. The outcome status was unknown for 2 cases (0.3%) (S2 Table). The eight expired cases were elderly (72-91 years old; 1 male and 7 females) and 2 cases were middle-aged adults (44 and 54 years old; both males). Seven cases had comorbidities involving multiple organ systems whilst three suffered from hypertension and the remaining one had an underlying malignancy. Respiratory failure was the most common cause of death (Seven out of ten deaths).

### Pediatric (0-19 years) reinfection cases

Out of 577 cases, disaggregated data for 24 pediatric (0-18 years) cases was available. Disaggregation reported positive contact histories in 7 cases (29.2%) with only 1 (4.2%) reporting a comorbidity. A total of 7 cases (29.2%) were asymptomatic followed by fever (n= 4, 16.7%) and cough (n= 3, 12.5%) in first infection whereas during reinfection, asymptomatic presentation (n= 7, 29.2%) was followed by cough (n= 4, 16.7%) and then fever (n= 3, 12.5%). With 9 abnormal chest X-ray findings (37.5%), the most frequently used management modalities during first infection and reinfection were anti-viral (n=12) and traditional Chinese Medicine (n=12), respectively. All patient outcomes were reported as recovered.

### Quality assessment of included studies

Seventy-two studies were determined to be of good quality while nine studies were of fair quality (S3 Table). Studies were primarily downgraded for unclear study objectives[63], incomplete case definition[16, 28, 48, 66, 67, 70, 77, 83], non-consecutive subject recruitment[19, 27, 30, 34, 38, 39, 44–46, 49, 71, 79, 80, 86, 90], incomparable subjects[79], inadequate length of follow-up[43, 63], inadequate description of statistical methods[39, 46, 80] and inadequate description of results[39, 78]. The most common cause for downgrading studies was non-consecutive recruitment which raised concerns that the included sample could be biased towards a more severe presentation or included more individuals undergoing routine screening.

## Discussion

Both the developing and developed world are still battling the spread of COVID-19. A major concern that needs to be addressed is the appearance of reinfections in previously recovered COVID-19 patients.

Our review is the first, and largest, systematic review covering COVID-19 reinfection cases from over 22 countries, raising questions concerning vaccination and exploring a specific set of determinants that can facilitate reinfection in recovered individuals. Similar systematic reviews on COVID-19 reinfection have been conducted previously[92, 93] but none of those studies, or any conducted so far in the literature, have been as extensive as this review in terms of analyzing the clinical information between first infection and reinfection whilst covering a wide range of international and regional databases. One of the major strengths of this review is the substantial timeframe that it covers: January 2020 to March 2021, spanning a total of 81 studies with a widespread distribution of High-Income Countries (HICs) and LMICs to differentiate features of reinfection cases as per different settings. In addition, adult cases were separated from pediatric cases to differentiate between clinical features and identify the optimal treatment management strategies as per varying age groups. Furthermore, case reports and case series included in our study were quality assessed with 72 out of 81 studies reported to be of good quality. An analysis of only pediatric reinfection cases was also conducted in this review. Good prognosis and lower morbidity were reported in the pediatric cases, similar to the general COVID-19 disease course in the pediatric population[94]. Therefore, we suggest public health campaigns targeting people of younger age as they are at similar risk of reinfection as adults, to ensure elimination of complacency and enforcement of protective measures, such as face masks and social distancing.

A broad distribution was seen amongst severity of first infection compared to reinfection as well as management and number of symptomatic cases. The most reported clinical symptoms in our review were fever (41.4%) and cough (34.8%) in first infection with a frequency of 36.4% cases with fever and 34.8% cases with cough in reinfection, respectively. These findings are similar to a trend observed in the review on reinfections by Gidari et al.[95] In addition, the number of asymptomatic cases in our review increased from 9.2% in first infection to 31.9% in second infection, similar to findings reported in the review by Gidari et al[95]. On the contrary, a higher requirement of ICU admission and mechanical ventilation was observed during reinfection in our review. A meta-analysis analysis of 123 cases by Vancsa et al[96] showed that the second episode of SARS-COV-2 infection is more severe than the first if it happens within 60 days of the first positive PCR. This deems the necessity of IPD analysis as many of the larger case series report more severe cases which might skew the overall findings. A total of 10 deaths were reported in this review, all amongst reinfection cases. In all 10 cases, several comorbidities were present and all patients who were classified as the most severe were of older age, a similar trend seen in a study by Wang et al.[97] Most of these patients died due to respiratory complications; similar effects of these comorbidities can be seen in other respiratory illnesses such as MERS-CoV[98]. The results from this review suggest that comorbidities and age play a major role in the outcome of critical patients.

The time duration between first infection and reinfection has been a source of debate. Alinaghi et al. in their systematic review[93] estimated that antibodies from natural infection lasted 40 days, after which the chances of reinfection increased. The average time duration between first infection and reinfection in our review was 63.6 days. Wang et al. in their review noted it to be 76 days[99] whereas Manish et al. in their review[100] observed a lengthier duration of median 113.5 days. This variation in time duration outlines the need for vigilance when it comes to COVID-19 reinfection, especially considering waning antibodies.

Another observation made in our review was differences in the presence of antibodies during first infection and reinfection. A recently published systematic review looked at antibody response following SARS-COV-2 infection across multiple studies[101]. They noted that 80% of patients developed IgM antibodies with antibodies being detected after a mean period of 7 days and declining after 27 days, and 95% of patients developed IgG antibodies with antibodies being detectable after a mean period of 12 days and started declining after 60 days. Likewise, IgA levels and neutralizing antibodies started declining after 30 days. In our review, 56.0% and 63.0% of the patients detected positive for antibodies during first infection and reinfection, respectively. Some evidence suggests that waning antibodies places individuals at a risk for reinfection, which may explain our finding of the time duration of first infection and reinfection being a mean of 63 days. The presence of antibodies could provide a protective role, but it does not specifically prevent reinfection as supported by findings in a systematic review by Piri et al.[102]

Whilst our review did not analyze the cause of reinfection being due to different variants, they cannot be excluded. A review by Wang et al.[99] concluded that previous COVID-19 reinfection did not confer total immunity and a second infection by a different variant was possible with the second infection being more severe than the first. Even though most of the studies in our review predated the announcements of the new variants, however, given the ability of the virus to mutate at a rapid pace, some reinfection cases reported in our study could be due to the variants which would have resulted in more severity of reinfection. Whether waning antibodies or new variants were the source of reinfection is a question that should be explored further in future studies.

A recent systematic review by Azam et al. looked at the incidence of SARS-COV-2 positivity in patients who had recovered from COVID-19 [103]. They noted that younger patients and those with a longer initial infection were more likely to have recurrent positivity. A similar systematic review by Manish et al. on the assessment of SARS-COV-2 mutations in reinfections and persistent infections[100] noted it to be challenging to differentiate between reinfection and persistent recurrent infection, concluding that the former happened in immunocompetent individuals and the latter happened in immunocompromised individuals. Furthermore, this phenomenon was associated with a faster viral evolution and mutation resulting in the creation of new variants. Lastly, another systematic review by Hoang on the risk factors associated with re-positive viral RNA after recovery from COVID-19 [104] postulated that the re-positive viral RNA seen in their review likely added to the evidence that viral relapse was a cause of COVID-19 recurrence.

This review has some limitations, such as the small sample sizes analyzed from each country except for China that had 73% (n=423) of the total included cases. The majority of these cases were reported from Wuhan or the Hubei province, where the gross domestic product per capita is less than half of that of Beijing and Shanghai[105]. Therefore, the findings of studies from China may be generalizable to the socioeconomic and health development status of other middle-income countries and not to high-income nations. This review can be improved by sampling larger series and including IPD, if available, to predict the outcome of COVID-19 illness based off epidemiological trends dramatically reducing hospitalization time, given the lack of sufficient healthcare resources in low-middle income countries. Therefore, a selection bias remains when considering LMICs where admitted hospital patients could be in a more critical state reporting a higher mortality rate.

Our review on reinfections in COVID-19 also comes at a pertinent time as countries, especially the developing world, suffer a repeated wave of infection[106]. At this time, public health initiatives aimed at removing complacency are the need of hour, and one of the key messages that needs to be given is that reinfection is a reality and vaccines along with social distancing remain the key in fighting the pandemic. A recently published online longitudinal survey [107] in 23 countries of high, middle and low income, across 4 continents with over 1 million participants provides hope in this regard, as it identified that the intention to vaccinate amongst the general public is at an all-time high, with the major issue not being vaccine hesitancy but a shortage of vaccines. A recent report of a 4 month surveillance of mass immunization in Israel [108] notes two doses of the Pfizer BioNTech mRNA COVId-19 vaccine to be highly effective (95.3%; 95% CI 94·9–95·7) against SARS-CoV-2 infection and mitigated COVID-19-related hospitalizations, severe disease, and death, including those caused by variants including the B.1.1.7 SARS-CoV-2. Whilst the world is still under process of getting vaccinated, data needs to be collected on patients in the long run to analyze whether vaccination has any correlation with reinfection cases and further investigate the average time needed by the various vaccines to achieve their desired efficacies. We hope that governments across the world seize this moment and take steps to ensure equitable distribution of vaccines so that the world can finally step out of the long shadow cast by the COVID-19 pandemic.

## Conclusion

COVID-19 first infections and reinfections observe a similar clinical spectrum and management regimen with a slightly higher severity reported during reinfection in the form of requirement for mechanical ventilation and ICU admission. There lies a need for much closer scrutiny of reinfections globally with individual patient data analysis to derive determinants of reinfection incidence and disposition to a severe infection.

## Supporting information

S1 Appendix. PRISMA checklist

S1 Table. Search Strategy for MEDLINE

S1 Fig. Global map of distribution of the studies included (N=81) in the review

S2 Table. Characteristics of included studies (N=81)

S3 Table. Quality assessment of case reports and case series (N=81)

## Data Availability

All data generated or analysed during this study are included in this article and its supporting information files. Further enquiries can be directed to the corresponding author.

## Acknowledgements

None.

## Supporting information

**S1 Table. Search Strategy for MEDLINE**

**S2 Table. Characteristics of included studies (N=81)**

**S3 Table. Quality assessment of case reports and case series (N=81)**

**S1 Fig. Global map of distribution of the studies included (N=81) in the review**

**S1 Appendix. PRISMA checklist**

## References

1. COVID-19 Dashboard [Internet]. Johns Hopkins University. 2021 [cited 20th May 2021]. Available from: https://coronavirus.jhu.edu/map.html.

2. Fergie J, Srivastava A. Immunity to SARS-CoV-2: Lessons Learned. Front Immunol. 2021;12:654165. Epub 2021/04/06. doi: 10.3389/fimmu.2021.654165. PubMed PMID: 33815415; PubMed Central PMCID: PMCPMC8018176.

3. Costa AOC, Neto HdCA, Nunes APL, de Castro RD, de Almeida RN. COVID-19: Is reinfection possible? EXCLI Journal. 2021;20:522–36.

4. Khoshkam Z, Aftabi Y, Stenvinkel P, Paige Lawrence B, Rezaei MH, Ichihara G, et al. Recovery scenario and immunity in COVID-19 disease: A new strategy to predict the potential of reinfection. J Adv Res. 2021. Epub 2021/02/02. doi: 10.1016/j.jare.2020.12.013. PubMed PMID: 33520309; PubMed Central PMCID: PMCPMC7832464.

5. Prevost J, Finzi A. The great escape? SARS-CoV-2 variants evading neutralizing responses. Cell Host Microbe. 2021;29(3):322-4. Epub 2021/03/12. doi: 10.1016/j.chom.2021.02.010. PubMed PMID: 33705702; PubMed Central PMCID: PMCPMC7945862.

6. Wang Z, Schmidt F, Weisblum Y, Muecksch F, Barnes CO, Finkin S, et al. mRNA vaccine-elicited antibodies to SARS-CoV-2 and circulating variants. Nature. 2021. Epub 2021/02/11. doi: 10.1038/s41586-021-03324-6. PubMed PMID: 33567448.

7. CDC. Reinfection with COVID-19. https://www.cdc.gov/coronavirus/2019-ncov/your-health/reinfection.html.2020.

8. Tillett RL, Sevinsky JR, Hartley PD, Kerwin H, Crawford N, Gorzalski A, et al. Genomic evidence for reinfection with SARS-CoV-2: a case study. Lancet Infect Dis. 2021;21(1):52–8. Epub 2020/10/16. doi: 10.1016/S1473-3099(20)30764-7. PubMed PMID: 33058797; PubMed Central PMCID: PMCPMC7550103.

9. Iwasaki A. What reinfections mean for COVID-19. The Lancet Infectious Diseases. 2021;21(1):3–5. doi: 10.1016/s1473-3099(20)30783-0.

10. Gomez CE, Perdiguero B, Esteban M. Emerging SARS-CoV-2 Variants and Impact in Global Vaccination Programs against SARS-CoV-2/COVID-19. Vaccines (Basel). 2021;9(3). Epub 2021/04/04. doi: 10.3390/vaccines9030243. PubMed PMID: 33799505; PubMed Central PMCID: PMCPMC7999234.

11. Welfare MoHAF. Genome Sequencing by INSACOG shows variants of concern and a Novel variant in India. https://pib.gov.in/PressReleaseIframePage.aspx?PRID=1707177: PIB Delhi; 2021.

12. Abdallah H, Porterfield F, Fajgenbaum D. Symptomatic relapse and long-term sequelae of COVID-19 in a previously healthy 30-year-old man. BMJ Case Rep. 2020;13(12):e239825-e. Epub 2020/12/16. doi: 10.1136/bcr-2020-239825. PubMed PMID: 33318288; PubMed Central PMCID: PMCPMC7736956.

13. Adrielle Dos Santos L, Filho PGG, Silva AMF, Santos JVG, Santos DS, Aquino MM, et al. Recurrent COVID-19 including evidence of reinfection and enhanced severity in thirty Brazilian healthcare workers. J Infect. 2021;82(3):399–406. Epub 2021/02/17. doi: 10.1016/j.jinf.2021.01.020. PubMed PMID: 33589297; PubMed Central PMCID: PMCPMC7880834.

14. Ak R, Yilmaz E, Seyhan AU, Doganay F. Recurrence of COVID-19 Documented with RT-PCR. J Coll Physicians Surg Pak. 2021;30(1):S26–S8. Epub 2021/03/03. doi: 10.29271/jcpsp.2021.01.S26. PubMed PMID: 33650420.

15. Ali AM, Ali KM, Fatah MH, Tawfeeq HM, Rostam HM. SARS-CoV-2 Reinfection in Patients Negative for Immunoglobulin G Following Recovery from COVID-19. medRxiv; 2020. p. 2020.11.20.20234385-2020.11.20.

16. Alonso FOM, Sabino BD, Guimaraes M, Varella RB. Recurrence of SARS-CoV-2 infection with a more severe case after mild COVID-19, reversion of RT-qPCR for positive and late antibody response: Case report. J Med Virol. 2021;93(2):655–6. Epub 2020/08/17. doi: 10.1002/jmv.26432. PubMed PMID: 32797634; PubMed Central PMCID: PMCPMC7436374.

17. An J, Liao X, Xiao T, Qian S, Yuan J, Ye H, et al. Clinical characteristics of recovered COVID-19 patients with re-detectable positive RNA test. Ann Transl Med. 2020;8(17):1084. Epub 2020/11/05. doi: 10.21037/atm-20-5602. PubMed PMID: 33145303; PubMed Central PMCID: PMCPMC7575971.

18. Arteaga-Livias K, Panduro-Correa V, Pinzas-Acosta K, Perez-Abad L, Pecho-Silva S, Espinoza-Sanchez F, et al. COVID-19 reinfection? A suspected case in a Peruvian patient. Travel Med Infect Dis. 2021;39(Jan-Feb 2021):101947. Epub 2020/12/12. doi: 10.1016/j.tmaid.2020.101947. PubMed PMID: 33307196; PubMed Central PMCID: PMCPMC7723440.

19. Atici S, Ek OF, Yildiz MS, Sikgenc MM, Guzel E, Soysal A. Symptomatic recurrence of SARS-CoV-2 infection in healthcare workers recovered from COVID-19. J Infect Dev Ctries. 2021;15(1):69–72. Epub 2021/02/12. doi: 10.3855/jidc.14305. PubMed PMID: 33571147.

20. Bellanti F, Lo Buglio A, Custodero G, Barbera L, Minafra G, Montrano M, et al. Fatal relapse of COVID-19 after recovery? A case report of an older Italian patient. J Infect. 2021;82(1):e49-e51. Epub 2020/12/20. doi: 10.1016/j.jinf.2020.12.009. PubMed PMID: 33340595; PubMed Central PMCID: PMCPMC7834308.

21. Bellesso M, Bruniera FR, Trunkel AT, Nicodemo IP. Second COVID-19 infection in a patient with multiple myeloma in Brazil - reinfection or reactivation? Hematol Transfus Cell Ther. 2021;43(1):109–11. Epub 2021/01/12. doi: 10.1016/j.htct.2020.12.002. PubMed PMID: 33423984; PubMed Central PMCID: PMCPMC7837121.

22. Bongiovanni M. COVID-19 reinfection in a healthcare worker. J Med Virol. 2020:jmv.26565-jmv. Epub 2020/09/30. doi: 10.1002/jmv.26565. PubMed PMID: 32990954; PubMed Central PMCID: PMCPMC7537129.

23. Bonifacio LP, Pereira APS, Araujo D, Balbao V, Fonseca B, Passos ADC, et al. Are SARS-CoV-2 reinfection and Covid-19 recurrence possible? a case report from Brazil. Rev Soc Bras Med Trop. 2020;53:e20200619. Epub 2020/09/24. doi: 10.1590/0037-8682-0619-2020. PubMed PMID: 32965458; PubMed Central PMCID: PMCPMC7508196.

24. Cao H, Ruan L, Liu J, Liao W. The clinical characteristic of eight patients of COVID-19 with positive RT-PCR test after discharge. J Med Virol. 2020;92(10):2159–64. Epub 2020/05/16. doi: 10.1002/jmv.26017. PubMed PMID: 32410245; PubMed Central PMCID: PMCPMC7272974.

25. Chan PKS, Lui G, Hachim A, Ko RLW, Boon SS, Li T, et al. Serologic Responses in Healthy Adult with SARS-CoV-2 Reinfection, Hong Kong, August 2020. Emerg Infect Dis. 2020;26(12):3076-8. Epub 2020/10/23. doi: 10.3201/eid2612.203833. PubMed PMID: 33089772; PubMed Central PMCID: PMCPMC7706979.

26. Chen D, Xu W, Lei Z, Huang Z, Liu J, Gao Z, et al. Recurrence of positive SARS-CoV-2 RNA in COVID-19: A case report. Int J Infect Dis. 2020;93(Apr):297–9. Epub 2020/03/10. doi: 10.1016/j.ijid.2020.03.003. PubMed PMID: 32147538; PubMed Central PMCID: PMCPMC7129213.

27. Chen J, Xu X, Hu J, Chen Q, Xu F, Liang H, et al. Clinical course and risk factors for recurrence of positive SARS-CoV-2 RNA: a retrospective cohort study from Wuhan, China. Aging (Albany NY). 2020;12(17):16675–89. Epub 2020/09/11. doi: 10.18632/aging.103795. PubMed PMID: 32909961; PubMed Central PMCID: PMCPMC7521537.

28. Colson P, Finaud M, Levy N, Lagier JC, Raoult D. Evidence of SARS-CoV-2 re-infection with a different genotype. J Infect. 2021;82(4):84–123. Epub 2020/11/19. doi: 10.1016/j.jinf.2020.11.011. PubMed PMID: 33207255; PubMed Central PMCID: PMCPMC7666873.

29. Coppola A, Annunziata A, Carannante N, Di Spirito V, Fiorentino G. Late Reactivation of SARS-CoV-2: A Case Report. Front Med (Lausanne). 2020;7:531. Epub 2020/09/26. doi: 10.3389/fmed.2020.00531. PubMed PMID: 32974374; PubMed Central PMCID: PMCPMC7468504.

30. de Brito CAA, Lima PMA, de Brito MCM, de Oliveira DB. Second Episode of COVID-19 in Health Professionals: Report of Two Cases. Int Med Case Rep J. 2020;13(Oct):471–5. Epub 2020/10/17. doi: 10.2147/IMCRJ.S277882. PubMed PMID: 33061670; PubMed Central PMCID: PMCPMC7537988.

31. Dou C, Xie X, Peng Z, Tang H, Jiang Z, Zhong Z, et al. A case presentation for positive SARS-CoV-2 RNA recurrence in a patient with a history of type 2 diabetes that had recovered from severe COVID-19. Diabetes Res Clin Pract. 2020;166:108300. Epub 2020/07/15. doi: 10.1016/j.diabres.2020.108300. PubMed PMID: 32663490; PubMed Central PMCID: PMCPMC7354258.

32. Du HW, Chen JN, Pan XB, Chen XL, Yixian Z, Fang SF, et al. Prevalence and outcomes of re-positive nucleic acid tests in discharged COVID-19 patients. Eur J Clin Microbiol Infect Dis. 2021;40(2):413–7. Epub 2020/09/01. doi: 10.1007/s10096-020-04024-1. PubMed PMID: 32865669; PubMed Central PMCID: PMCPMC7456660.

33. Duggan NM, Ludy SM, Shannon BC, Reisner AT, Wilcox SR. Is novel coronavirus 2019 reinfection possible? Interpreting dynamic SARS-CoV-2 test results. Am J Emerg Med. 2021;39:256 e1-e3. Epub 2020/07/25. doi: 10.1016/j.ajem.2020.06.079. PubMed PMID: 32703607; PubMed Central PMCID: PMCPMC7335242.

34. Fu W, Chen Q, Wang T. Letter to the Editor: Three cases of redetectable positive SARS-CoV-2 RNA in recovered COVID-19 patients with antibodies. J Med Virol. 2020;92(11):2298–301. Epub 2020/05/06. doi: 10.1002/jmv.25968. PubMed PMID: 32369214; PubMed Central PMCID: PMCPMC7267393.

35. Gao G, Zhu Z, Fan L, Ye S, Huang Z, Shi Q, et al. Absent immune response to SARS-CoV-2 in a 3-month recurrence of coronavirus disease 2019 (COVID-19) case. Infection. 2021;49(1):57–61. Epub 2020/07/30. doi: 10.1007/s15010-020-01485-6. PubMed PMID: 32725596; PubMed Central PMCID: PMCPMC7386381.

36. Goldman JD, Wang K, Röltgen K, Nielsen SCA, Roach JC, Naccache SN, et al. Reinfection with SARS-CoV-2 and Failure of Humoral Immunity: A case report. medRxiv; 2020.

37. Gousseff M, Penot P, Gallay L, Batisse D, Benech N, Bouiller K, et al. Clinical recurrences of COVID-19 symptoms after recovery: Viral relapse, reinfection or inflammatory rebound? J Infect. 2020;81(5):816–46. Epub 2020/07/04. doi: 10.1016/j.jinf.2020.06.073. PubMed PMID: 32619697; PubMed Central PMCID: PMCPMC7326402 interest to declare regarding this subject. This work had no financial support.

38. Gupta V, Bhoyar RC, Jain A, Srivastava S, Upadhayay R, Imran M, et al. Asymptomatic reinfection in two healthcare workers from India with genetically distinct SARS-CoV-2. Clin Infect Dis. 2020. Epub 2020/09/24. doi: 10.1093/cid/ciaa1451. PubMed PMID: 32964927; PubMed Central PMCID: PMCPMC7543380.

39. Habibzadeh P, Sajadi MM, Emami A, Karimi MH, Yadollahie M, Kucheki M, et al. Rate of re-positive RT-PCR test among patients recovered from COVID-19. Biochem Med (Zagreb). 2020;30(3):030401. Epub 2020/08/11. doi: 10.11613/BM.2020.030401. PubMed PMID: 32774117; PubMed Central PMCID: PMCPMC7394260.

40. Hanif M, Haider MA, Ali MJ, Naz S, Sundas F. Reinfection of COVID-19 in Pakistan: A First Case Report. Cureus. 2020;12(10):e11176. Epub 2020/12/03. doi: 10.7759/cureus.11176. PubMed PMID: 33262913; PubMed Central PMCID: PMCPMC7689968.

41. Harrington D, Kele B, Pereira S, Couto-Parada X, Riddell A, Forbes S, et al. Confirmed Reinfection with SARS-CoV-2 Variant VOC-202012/01. Clin Infect Dis. 2021. Epub 2021/01/10. doi: 10.1093/cid/ciab014. PubMed PMID: 33421056; PubMed Central PMCID: PMCPMC7929017.

42. He S, Sun W, Zhou K, Hu M, Liu C, Xie L, et al. Clinical Characteristics Analysis of the “Re-positive” Discharged COVID-19 Pneumonia Patients in Wuhan, China. 2020. doi: 10.21203/rs.3.rs-28667/v1.

43. Huang J, Zheng L, Li Z, Hao S, Ye F, Chen J, et al. Recurrence of SARS-CoV-2 PCR positivity in COVID-19 patients: A single center experience and potential implications. medRxiv; 2020. p. 2020.05.06.20089573-2020.05.06.

44. Kapoor R, Nair RK, Nayan N, Bhalla S, Singh J. Reinfection or Reactivation of Coronavirus-19 in Patients with Hematologic Malignancies: Case Report Series. SN Compr Clin Med. 2021;3(2):1–5. Epub 2021/02/16. doi: 10.1007/s42399-021-00790-x. PubMed PMID: 33585797; PubMed Central PMCID: PMCPMC7873512.

45. Lafaie L, Celarier T, Goethals L, Pozzetto B, Grange S, Ojardias E, et al. Recurrence or Relapse of COVID-19 in Older Patients: A Description of Three Cases. J Am Geriatr Soc. 2020;68(10):2179–83. Epub 2020/07/09. doi: 10.1111/jgs.16728. PubMed PMID: 32638347; PubMed Central PMCID: PMCPMC7361461.

46. Lan L, Xu D, Ye G, Xia C, Wang S, Li Y, et al. Positive RT-PCR Test Results in Patients Recovered From COVID-19. JAMA. 2020;323(15):1502–3. Epub 2020/02/28. doi: 10.1001/jama.2020.2783. PubMed PMID: 32105304; PubMed Central PMCID: PMCPMC7047852.

47. Lancman G, Mascarenhas J, Bar-Natan M. Severe COVID-19 virus reactivation following treatment for B cell acute lymphoblastic leukemia. J Hematol Oncol. 2020;13(1):131. Epub 2020/10/04. doi: 10.1186/s13045-020-00968-1. PubMed PMID: 33008453; PubMed Central PMCID: PMCPMC7531062.

48. Larson D, Brodniak SL, Voegtly LJ, Cer RZ, Glang LA, Malagon FJ, et al. A Case of Early Re-infection with SARS-CoV-2. Clin Infect Dis. 2020. Epub 2020/09/20. doi: 10.1093/cid/ciaa1436. PubMed PMID: 32949240; PubMed Central PMCID: PMCPMC7543357.

49. Li J, Wei X, Tian W, Zou J, Wang Y, Xue W, et al. Clinical features of discharged COVID-19 patients with an extended SARS-CoV-2 RNA positive signal in respiratory samples. Virus Res. 2020;286:198047. Epub 2020/06/12. doi: 10.1016/j.virusres.2020.198047. PubMed PMID: 32522535; PubMed Central PMCID: PMCPMC7833058.

50. Li XJ, Zhang ZW, Zong ZY. A case of a readmitted patient who recovered from COVID-19 in Chengdu, China. Crit Care. 2020;24(1):152. Epub 2020/04/18. doi: 10.1186/s13054-020-02877-8. PubMed PMID: 32299477; PubMed Central PMCID: PMCPMC7160612.

51. Li Y, Hu Y, Yu Y, Zhang X, Li B, Wu J, et al. Positive result of Sars-Cov-2 in faeces and sputum from discharged patients with COVID-19 in Yiwu, China. J Med Virol. 2020;92(10):1938–47. Epub 2020/04/21. doi: 10.1002/jmv.25905. PubMed PMID: 32311109; PubMed Central PMCID: PMCPMC7264799.

52. Ling Y, Xu SB, Lin YX, Tian D, Zhu ZQ, Dai FH, et al. Persistence and clearance of viral RNA in 2019 novel coronavirus disease rehabilitation patients. Chin Med J (Engl). 2020;133(9):1039–43. Epub 2020/03/03. doi: 10.1097/CM9.0000000000000774. PubMed PMID: 32118639; PubMed Central PMCID: PMCPMC7147278.

53. Liu F, Cai ZB, Huang JS, Yu WY, Niu HY, Zhang Y, et al. Positive SARS-CoV-2 RNA recurs repeatedly in a case recovered from COVID-19: dynamic results from 108 days of follow-up. Pathog Dis. 2020;78(4):31-. Epub 2020/06/28. doi: 10.1093/femspd/ftaa031. PubMed PMID: 32592396; PubMed Central PMCID: PMCPMC7337794.

54. Loconsole D, Passerini F, Palmieri VO, Centrone F, Sallustio A, Pugliese S, et al. Recurrence of COVID-19 after recovery: a case report from Italy. Infection. 2020;48(6):965–7. Epub 2020/05/18. doi: 10.1007/s15010-020-01444-1. PubMed PMID: 32415334; PubMed Central PMCID: PMCPMC7228864.

55. Luo A. Positive SARS-Cov-2 test in a woman with COVID-19 at 22 days after hospital discharge: A case report. Journal of Traditional Chinese Medical Sciences. 2020;7(4):413–7. doi: 10.1016/j.jtcms.2020.04.001.

56. Mei Q, Li J, Du R, Yuan X, Li M, Li J. Assessment of patients who tested positive for COVID-19 after recovery. Lancet Infect Dis. 2020;20(9):1004–5. Epub 2020/07/10. doi: 10.1016/S1473-3099(20)30433-3. PubMed PMID: 32645295; PubMed Central PMCID: PMCPMC7338012.

57. Moore JL, Ganapathiraju PV, Kurtz CP, Wainscoat B. A 63-Year-Old Woman with a History of Non-Hodgkin Lymphoma with Persistent SARS-CoV-2 Infection Who Was Seronegative and Treated with Convalescent Plasma. Am J Case Rep. 2020;21:e927812. Epub 2020/10/04. doi: 10.12659/AJCR.927812. PubMed PMID: 33009361; PubMed Central PMCID: PMCPMC7542548.

58. Mulder M, van der Vegt D, Oude Munnink BB, GeurtsvanKessel CH, van de Bovenkamp J, Sikkema RS, et al. Reinfection of SARS-CoV-2 in an immunocompromised patient: a case report. Clin Infect Dis. 2020. Epub 2020/10/13. doi: 10.1093/cid/ciaa1538. PubMed PMID: 33043962; PubMed Central PMCID: PMCPMC7665355.

59. Nachmias V, Fusman R, Mann S, Koren G. The first case of documented Covid-19 reinfection in Israel. IDCases. 2020;22:e00970. Epub 2020/10/09. doi: 10.1016/j.idcr.2020.e00970. PubMed PMID: 33029476; PubMed Central PMCID: PMCPMC7528892.

60. Novoa W, Miller H, Mattar S, Faccini-Martinez AA, Rivero R, Serrano-Coll H. A first probable case of SARS-CoV-2 reinfection in Colombia. Ann Clin Microbiol Antimicrob. 2021;20(1):7. Epub 2021/01/14. doi: 10.1186/s12941-020-00413-8. PubMed PMID: 33435982; PubMed Central PMCID: PMCPMC7802059.

61. Pan L, Wang R, Yu N, Hu C, Yan J, Zhang X, et al. Clinical characteristics of re-hospitalized COVID-19 patients with recurrent positive SARS-CoV-2 RNA: a retrospective study. Eur J Clin Microbiol Infect Dis. 2021. Epub 2021/01/16. doi: 10.1007/s10096-020-04151-9. PubMed PMID: 33447913; PubMed Central PMCID: PMCPMC7808928.

62. Parry J. Covid-19: Hong Kong scientists report first confirmed case of reinfection. BMJ. 2020;370:m3340. Epub 2020/08/28. doi: 10.1136/bmj.m3340. PubMed PMID: 32847834.

63. Patrocinio de Jesus R, Silva R, Aliyeva E, Lopes L, Portugalyan M, Antunes L, et al. Reactivation of SARS-CoV-2 after Asymptomatic Infection while on High-Dose Corticosteroids. Case Report. SN Compr Clin Med. 2020;2(11):1–4. Epub 2020/10/13. doi: 10.1007/s42399-020-00548-x. PubMed PMID: 33043249; PubMed Central PMCID: PMCPMC7531809.

64. Prado-Vivar B, Becerra-Wong M, Guadalupe JJ, Marquez S, Gutierrez B, Rojas-Silva P, et al. A case of SARS-CoV-2 reinfection in Ecuador. Lancet Infect Dis. 2020. Epub 2020/11/27. doi: 10.1016/S1473-3099(20)30910-5. PubMed PMID: 33242475; PubMed Central PMCID: PMCPMC7833993.

65. Salcin S, Fontem F. Recurrent SARS-CoV-2 infection resulting in acute respiratory distress syndrome and development of pulmonary hypertension: A case report. Respir Med Case Rep. 2021;33:101314. Epub 2020/12/15. doi: 10.1016/j.rmcr.2020.101314. PubMed PMID: 33312856; PubMed Central PMCID: PMCPMC7718582.

66. Selhorst P, Van Ierssel S, Michiels J, Marien J, Bartholomeeusen K, Dirinck E, et al. Symptomatic SARS-CoV-2 reinfection of a health care worker in a Belgian nosocomial outbreak despite primary neutralizing antibody response. Clin Infect Dis. 2020. Epub 2020/12/15. doi: 10.1093/cid/ciaa1850. PubMed PMID: 33315049; PubMed Central PMCID: PMCPMC7799230.

67. Selvaraj V, Herman K, Dapaah-Afriyie K. Severe, Symptomatic Reinfection in a Patient with COVID-19. R I Med J (2013). 2020;103(10):24-6. Epub 2020/11/12. PubMed PMID: 33172223.

68. Sharma R, Sardar S, Mohammad Arshad A, Ata F, Zara S, Munir W. A Patient with Asymptomatic SARS-CoV-2 Infection Who Presented 86 Days Later with COVID-19 Pneumonia Possibly Due to Reinfection with SARS-CoV-2. Am J Case Rep. 2020;21:e927154. Epub 2020/12/02. doi: 10.12659/AJCR.927154. PubMed PMID: 33257644; PubMed Central PMCID: PMCPMC7718490.

69. Sicsic I, Jr., Chacon AR, Zaw M, Ascher K, Abreu A, Chediak A. A case of SARS-CoV-2 reinfection in a patient with obstructive sleep apnea managed with telemedicine. BMJ Case Rep. 2021;14(2):240496-. Epub 2021/02/03. doi: 10.1136/bcr-2020-240496. PubMed PMID: 33526540; PubMed Central PMCID: PMCPMC7852971.

70. Song KH, Kim DM, Lee H, Ham SY, Oh SM, Jeong H, et al. Dynamics of viral load and anti-SARS-CoV-2 antibodies in patients with positive RT-PCR results after recovery from COVID-19. Korean J Intern Med. 2021;36(1):11–4. Epub 2020/09/26. doi: 10.3904/kjim.2020.325. PubMed PMID: 32972123; PubMed Central PMCID: PMCPMC7820639.

71. Fernandes Valente Takeda C, Moura de Almeida M, Goncalves de Aguiar Gomes R, Cisne Souza T, Alves de Lima Mota M, Pamplona de Goes Cavalcanti L, et al. Case Report: Recurrent Clinical Symptoms of COVID-19 in Healthcare Professionals: A Series of Cases from Brazil. Am J Trop Med Hyg. 2020;103(5):1993–6. Epub 2020/09/06. doi: 10.4269/ajtmh.20-0893. PubMed PMID: 32888288; PubMed Central PMCID: PMCPMC7646791.

72. Tian M, Long Y, Hong Y, Zhang X, Zha Y. The treatment and follow-up of ‘recurrence’ with discharged COVID-19 patients: data from Guizhou, China. Environ Microbiol. 2020;22(8):3588–92. Epub 2020/07/08. doi: 10.1111/1462-2920.15156. PubMed PMID: 32632947; PubMed Central PMCID: PMCPMC7361525.

73. To KK, Hung IF, Ip JD, Chu AW, Chan WM, Tam AR, et al. COVID-19 re-infection by a phylogenetically distinct SARS-coronavirus-2 strain confirmed by whole genome sequencing. Clin Infect Dis. 2020. Epub 2020/08/26. doi: 10.1093/cid/ciaa1275. PubMed PMID: 32840608; PubMed Central PMCID: PMCPMC7499500.

74. Tomassini S, Kotecha D, Bird PW, Folwell A, Biju S, Tang JW. Setting the criteria for SARS-CoV-2 reinfection - six possible cases. J Infect. 2021;82(2):282–327. Epub 2020/08/18. doi: 10.1016/j.jinf.2020.08.011. PubMed PMID: 32800801; PubMed Central PMCID: PMCPMC7422822.

75. Torres DA, Ribeiro L, Riello A, Horovitz DDG, Pinto LFR, Croda J. Reinfection of COVID-19 after 3 months with a distinct and more aggressive clinical presentation: Case report. J Med Virol. 2021;93(4):1857–9. Epub 2020/10/29. doi: 10.1002/jmv.26637. PubMed PMID: 33112002.

76. Van Elslande J, Vermeersch P, Vandervoort K, Wawina-Bokalanga T, Vanmechelen B, Wollants E, et al. Symptomatic SARS-CoV-2 reinfection by a phylogenetically distinct strain. Clin Infect Dis. 2020. Epub 2020/09/06. doi: 10.1093/cid/ciaa1330. PubMed PMID: 32887979; PubMed Central PMCID: PMCPMC7499557.

77. Vetter P, Cordey S, Schibler M, Vieux L, Despres L, Laubscher F, et al. Clinical, virologic and immunologic features of a mild case of SARS-CoV-2 reinfection. Clin Microbiol Infect. 2021;:1-. Epub 2021/02/23. doi: 10.1016/j.cmi.2021.02.010. PubMed PMID: 33618012; PubMed Central PMCID: PMCPMC7896115.

78. West J, Everden S, Nikitas N. A case of COVID-19 reinfection in the UK. Clin Med (Lond). 2021;21(1):e52–e3. Epub 2020/12/12. doi: 10.7861/clinmed.2020-0912. PubMed PMID: 33303623; PubMed Central PMCID: PMCPMC7850175.

79. Wu J, Cheng J, Shi X, Liu J, Huang B, Zhao X, et al. Recurrence of SARS-CoV-2 nucleic acid positive test in patients with COVID-19: a report of two cases. BMC Pulm Med. 2020;20(1):308. Epub 2020/11/24. doi: 10.1186/s12890-020-01348-8. PubMed PMID: 33225932; PubMed Central PMCID: PMCPMC7681189.

80. Yadav S, Wadhwa T, Thakkar D, Kapoor R, Rastogi N, Sarma S. Covid19 Reinfection in Two Children with Cancer. Authorea Preprints. 2020. doi: 10.22541/au.159986505.57940176.

81. Yadav SP, Thakkar D, Bhoyar RC, Jain A, Wadhwa T, Imran M, et al. Asymptomatic reactivation of SARS-CoV-2 in a child with neuroblastoma characterised by whole genome sequencing. IDCases. 2021;23:e01018. Epub 2020/12/09. doi: 10.1016/j.idcr.2020.e01018. PubMed PMID: 33288996; PubMed Central PMCID: PMCPMC7711173.

82. Ye G, Pan Z, Pan Y, Deng Q, Chen L, Li J, et al. Clinical characteristics of severe acute respiratory syndrome coronavirus 2 reactivation. J Infect. 2020;80(5):e14–e7. Epub 2020/03/17. doi: 10.1016/j.jinf.2020.03.001. PubMed PMID: 32171867; PubMed Central PMCID: PMCPMC7102560.

83. Yoo SY, Lee Y, Lee GH, Kim DH. Reactivation of SARS-CoV-2 after recovery. Pediatr Int. 2020;62(7):879–81. Epub 2020/05/19. doi: 10.1111/ped.14312. PubMed PMID: 32421910; PubMed Central PMCID: PMCPMC7276786.

84. Yuan J, Kou S, Liang Y, Zeng J, Pan Y, Liu L. Polymerase Chain Reaction Assays Reverted to Positive in 25 Discharged Patients With COVID-19. Clin Infect Dis. 2020;71(16):2230–2. Epub 2020/04/09. doi: 10.1093/cid/ciaa398. PubMed PMID: 32266381; PubMed Central PMCID: PMCPMC7184423.

85. Zayet S, Royer PY, Toko L, Pierron A, Gendrin V, Klopfenstein T. Recurrence of COVID-19 after recoveryA case series in health care workers, France. Microbes Infect. 2021:104803. Epub 2021/03/06. doi: 10.1016/j.micinf.2021.104803. PubMed PMID: 33667643; PubMed Central PMCID: PMCPMC7923857.

86. Zhang B, Liu S, Dong Y, Zhang L, Zhong Q, Zou Y, et al. Positive rectal swabs in young patients recovered from coronavirus disease 2019 (COVID-19). J Infect. 2020;81(2):e49–e52. Epub 2020/04/27. doi: 10.1016/j.jinf.2020.04.023. PubMed PMID: 32335176; PubMed Central PMCID: PMCPMC7177113.

87. Zhao W, Wang Y, Tang Y, Zhao W, Fan Y, Liu G, et al. Characteristics of Children With Reactivation of SARS-CoV-2 Infection After Hospital Discharge. Clin Pediatr (Phila). 2020;59(9-10):929–32. Epub 2020/05/29. doi: 10.1177/0009922820928057. PubMed PMID: 32462940.

88. Zheng J, Zhou R, Chen F, Tang G, Wu K, Li F, et al. Incidence, clinical course and risk factor for recurrent PCR positivity in discharged COVID-19 patients in Guangzhou, China: A prospective cohort study. PLoS Negl Trop Dis. 2020;14(8):e0008648. Epub 2020/09/01. doi: 10.1371/journal.pntd.0008648. PubMed PMID: 32866168; PubMed Central PMCID: PMCPMC7505432.

89. Zheng KI, Wang XB, Jin XH, Liu WY, Gao F, Chen YP, et al. A Case Series of Recurrent Viral RNA Positivity in Recovered COVID-19 Chinese Patients. J Gen Intern Med. 2020;35(7):2205–6. Epub 2020/04/22. doi: 10.1007/s11606-020-05822-1. PubMed PMID: 32314129; PubMed Central PMCID: PMCPMC7169645.

90. Zhu H, Fu L, Jin Y, Shao J, Zhang S, Zheng N, et al. Clinical features of COVID-19 convalescent patients with re-positive nucleic acid detection. J Clin Lab Anal. 2020;34(7):e23392. Epub 2020/06/09. doi: 10.1002/jcla.23392. PubMed PMID: 32506726; PubMed Central PMCID: PMCPMC7300578.

91. Zucman N, Uhel F, Descamps D, Roux D, Ricard JD. Severe reinfection with South African SARS-CoV-2 variant 501Y.V2: A case report. Clin Infect Dis. 2021. Epub 2021/02/11. doi: 10.1093/cid/ciab129. PubMed PMID: 33566076; PubMed Central PMCID: PMCPMC7929064.

92. Padda I, Khehra N, Jaferi U, Mosabbeh D, Atwal H, Musaji A, et al. Organ system effects and reinfection of COVID-19: A systematic review. Journal of Research in Clinical Medicine. 2021;9(1):6-. doi: 10.34172/jrcm.2021.006.

93. SeyedAlinaghi S, Oliaei S, Kianzad S, Afsahi AM, MohsseniPour M, Barzegary A, et al. Reinfection risk of novel coronavirus (COVID-19): A systematic review of current evidence. World J Virol. 2020;9(5):79–90. Epub 2020/12/29. doi: 10.5501/wjv.v9.i5.79. PubMed PMID: 33363000; PubMed Central PMCID: PMCPMC7747024.

94. Irfan O, Muttalib F, Tang K, Jiang L, Lassi ZS, Bhutta Z. Clinical characteristics, treatment and outcomes of paediatric COVID-19: a systematic review and meta-analysis. Arch Dis Child. 2021. Epub 2021/02/18. doi: 10.1136/archdischild-2020-321385. PubMed PMID: 33593743.

95. Gidari A, Nofri M, Saccarelli L, Bastianelli S, Sabbatini S, Bozza S, et al. Is recurrence possible in coronavirus disease 2019 (COVID-19)? Case series and systematic review of literature. Eur J Clin Microbiol Infect Dis. 2021;40(1):1-12. Epub 2020/10/11. doi: 10.1007/s10096-020-04057-6. PubMed PMID: 33037944; PubMed Central PMCID: PMCPMC7547550.

96. Vancsa S, Dembrovszky F, Farkas N, Szako L, Teutsch B, Bunduc S, et al. Repeated SARS-CoV-2 Positivity: Analysis of 123 Cases. Viruses. 2021;13(3). Epub 2021/04/04. doi: 10.3390/v13030512. PubMed PMID: 33808867; PubMed Central PMCID: PMCPMC8003803.

97. Wang D, Hu B, Hu C, Zhu F, Liu X, Zhang J, et al. Clinical Characteristics of 138 Hospitalized Patients With 2019 Novel Coronavirus-Infected Pneumonia in Wuhan, China. JAMA. 2020;323(11):1061–9. Epub 2020/02/08. doi: 10.1001/jama.2020.1585. PubMed PMID: 32031570; PubMed Central PMCID: PMCPMC7042881.

98. Badawi A, Ryoo SG. Prevalence of comorbidities in the Middle East respiratory syndrome coronavirus (MERS-CoV): a systematic review and meta-analysis. Int J Infect Dis. 2016;49(Aug):129–33. Epub 2016/06/30. doi: 10.1016/j.ijid.2016.06.015. PubMed PMID: 27352628; PubMed Central PMCID: PMCPMC7110556.

99. Wang J, Kaperak C, Sato T, Sakuraba A. COVID-19 reinfection: A Rapid Systematic Review of Case Reports and Case Series. medRxiv. 2021:2021.03.22.21254081. doi: 10.1101/2021.03.22.21254081.

100. Choudhary MC, Crain CR, Qiu X, Hanage W, Li JZ. SARS-CoV-2 Sequence Characteristics of COVID-19 Persistence and Reinfection. Clin Infect Dis. 2021;(2021). Epub 2021/04/28. doi: 10.1093/cid/ciab380. PubMed PMID: 33906227.

101. Arkhipova-Jenkins I, Helfand M, Armstrong C, Gean E, Anderson J, Paynter RA, et al. Antibody Response After SARS-CoV-2 Infection and Implications for Immunity : A Rapid Living Review. Ann Intern Med. 2021. Epub 2021/03/16. doi: 10.7326/M20-7547. PubMed PMID: 33721517; PubMed Central PMCID: PMCPMC8025942.

102. Piri SM, Edalatfar M, Shool S, Jalalian MN, Tavakolpour S. A systematic review on the recurrence of SARS-CoV-2 virus: frequency, risk factors, and possible explanations. Infect Dis (Lond). 2021;53(5):315–24. Epub 2021/01/30. doi: 10.1080/23744235.2020.1871066. PubMed PMID: 33508989; PubMed Central PMCID: PMCPMC7852280.

103. Azam M, Sulistiana R, Ratnawati M, Fibriana AI, Bahrudin U, Widyaningrum D, et al. Recurrent SARS-CoV-2 RNA positivity after COVID-19: a systematic review and meta-analysis. Sci Rep. 2020;10(1):20692. Epub 2020/11/28. doi: 10.1038/s41598-020-77739-y. PubMed PMID: 33244060; PubMed Central PMCID: PMCPMC7691365.

104. Hoang T. Systematic review and meta-analysis of factors associated with re-positive viral RNA after recovery from COVID-19. J Med Virol. 2021;93(4):2234–42. Epub 2020/11/03. doi: 10.1002/jmv.26648. PubMed PMID: 33135788.

105. NBS. Home - Regional - Quarterly by Province. https://data.stats.gov.cn/english/easyquery.htm?cn=E0102: National Bureau of Statistics, China 2019.

106. Gan N. Beyond India, a growing number of Asian countries are being ravaged by fresh coronavirus waves. CNN. 2021 May 8 2021.

107. Kothari A, Pfuhl G, Schieferdecker D, Harris CT, Tidwell C, Fitzpatrick KM, et al. The Barrier to Vaccination Is Not Vaccine Hesitancy: Patterns of COVID-19 Vaccine Acceptance over the Course of the Pandemic in 23 Countries. medRxiv. 2021:2021.04.23.21253857. doi: 10.1101/2021.04.23.21253857.

108. Haas EJ, Angulo FJ, McLaughlin JM, Anis E, Singer SR, Khan F, et al. Impact and effectiveness of mRNA BNT162b2 vaccine against SARS-CoV-2 infections and COVID-19 cases, hospitalisations, and deaths following a nationwide vaccination campaign in Israel: an observational study using national surveillance data. The Lancet. doi: 10.1016/S0140-6736(21)00947-8.

