## Supplementary material for "The mystery of COVID-19 reinfections: A global systematic review and meta-analysis of 577 cases": S1 Table. Search Strategy for MEDLINE

| | |
|---|---|
| **1** | (coronavirus or "covid 2019" or "SARS2" or "SARS" or "SARS CoV19" or "severe acute respiratory syndrome coronavirus 2" or "coronavirus infection" or "severe acute respiratory pneumonia outbreak" or "novel cov" or 2019ncov or sars cov2 or cov2 or ncov or covid or covid19 or coronaviridae or "corona virus").mp. [mp=title, abstract, original title, name of substance word, subject heading word, floating sub-heading word, keyword heading word, organism supplementary concept word, protocol supplementary concept word, rare disease supplementary concept word, unique identifier, synonyms] |
| **2** | "coronavirus infections".mp. or exp Coronavirus Infections/ |
| **3** | 1 or 2 |
| **4** | (“reinfection” or “reinfect*” or “reinfected” or “reinfecting” or “reinfections” or “reinfects” or “reinfestation” or “reinfest*” or “reinfested” or “relapse*” or “relapsed” or “relapsing” or “recurrent” or “recurrence” or “recontamination” or “recontaminate*” or “recontaminated”) mp. [mp=title, abstract, original title, name of substance word, subject heading word, floating sub-heading word, keyword heading word, organism supplementary concept word, protocol supplementary concept word, rare disease supplementary concept word, unique identifier, synonyms] |
| **5** | 3 and 4 |
