## Supplementary figures and images for "The mystery of COVID-19 reinfections: A global systematic review and meta-analysis of 577 cases"

### S1 Fig. Global map of distribution of the studies included (N=81) in the review

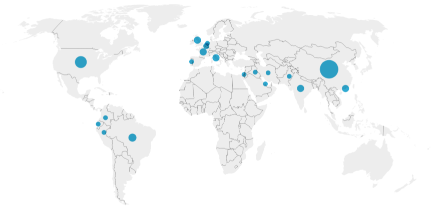
