## Supplementary material for "The mystery of COVID-19 reinfections: A global systematic review and meta-analysis of 577 cases": S2 Table. Characteristics of included studies (N=81)

| Study and year | Study Design | Country | World Bank Country Classification  LMIC: Low and middle-income Country, HIC: High-income Country | Total number (N) | Mean age (years) | Males to Female (N) | Past medical history and Contact history | Presenting signs and symptoms | Management (other than supportive care) | | Outcomes after reinfection |
| --- | --- | --- | --- | --- | --- | --- | --- | --- | --- | --- | --- |
| A Coppola et al, 2020 | Case report | Italy | HIC | 1 | 68 | 1 to 0 | Comorbidities (n=4) | First infection  Fever (n=1), diarrhea (n=1)  Reinfection  Fever (n=1), myalgia (n=1), diarrhea (n=1), fatigue (n=1) | | First infection  Steroids (n=1), antivirals (n=1), antibiotics (n=1), low flow oxygen (n=1)  Reinfection  Steroids (n=1), antivirals (n=1), antibiotics (n=1), low flow oxygen (n=1) | Recovered (n=1) |
| A Luo, 2020 | Case report | China | LMIC | 1 | 58 | 0 to 1 | Contact history (n=1), comorbidities (n=1) | First infection  Fever (n=1), cough (n=1), fatigue (n=1)  Reinfection  Cough (n=1), sore throat (n=1) | | First infection  Traditional medicine (n=1)  Reinfection  Traditional medicine (n=1) | Recovered (n=1) |
| AM Ali et al, 2020 | Case series | Iraq | LMIC | 26 | 35 | 14 to 12 | No contact history or comorbidities | First infection  Fever (n=25), cough (n=3), myalgia (n=18)  Reinfection  Fever (n=24), cough (n=22), myalgia (n=22), dyspnea (n=7), asymptomatic (n=1) | | First infection  Supportive care only  Reinfection  Supportive care only | Recovered (n=26) |
| B Prado-Vivar et al, 2020 | Case report | Ecuador | LMIC | 1 | 46 | 1 to 0 | No contact history or comorbidities | First infection  Headache (n=1),  Reinfection  Fever (n=1), sore throat (n=1), dyspnea (n=1) | | First infection  Supportive care only  Reinfection  Supportive care only | Recovered (n=1) |
| B Zhang et al, 2020 | Case series | China | LMIC | 7 | 18 | 6 to 1 | Contact history (n=7), comorbidities (n=1) | First infection  Fever (n=4), cough (n=3), fatigue (n=7)  Reinfection  Asymptomatic (n=7) | | First infection  Antivirals (n=7), interferon (n=4), low flow oxygen (n=6)  Reinfection  Traditional medicine (n=7) | Recovered (n=7) |
| C Dou et al, 2020 | Case report | China | LMIC | 1 | 34 | 1 to 0 | Comorbidities (n=1) | First infection  Fever (n=1), cough (n=1), fatigue (n=1), sore throat (n=1)  Reinfection  Asymptomatic (n=1) | | First infection  Antivirals (n=1), antibiotics (n=1)  Reinfection  Steroids (n=1), antivirals (n=1), traditional medicine (n=1), interferon (n=1) | Recovered (n=1) |
| CAA de Brito et al, 2020 | Case series | Brazil | LMIC | 2 | 43.5 | 1 to 1 | Contact History (n=2) | First infection  Fever (n=2), cough (n=1), myalgia (n=1), diarrhea (n=1), fatigue (n=2), sore throat (n=2), headache (n=1), dyspnea (n=1)  Reinfection  Fever(n=2), cough (n=1), myalgia (n=1), diarrhea (n=1), fatigue (n=2), sore throat (n=2), headache (n=1), dyspnea (n=1) | | First infection  Antivirals (n=2), antibiotics (n=2),  Reinfection  Antivirals (n=2), antibiotics (n=2) | Recovered (n=2) |
| CFV Takeda et al, 2020 | Case series | Brazil | LMIC | 6 | 46.2 | 5 to 1 | Comorbidities (n=5) | First infection  Fever (n=5), cough (n=3), myalgia (n=4), diarrhea (n=1), sore throat (n=2), headache (n=2), dyspnea (n=2)  Reinfection  Fever (n=4), fatigue (n=3), dyspnea (n=1) | | First infection  Steroids (n=4), antivirals (n=4), antibiotics (n=4)  Reinfection  Steroids (n=5), antivirals (n=5), antibiotics (n=5) | Recovered (n=6) |
| D Chen et al, 2020 | Case report | China | LMIC | 1 | 46 | 0 to 1 | Contact history (n=1) | First infection  Fever (n=1), cough (n=1), sore throat (n=1)  Reinfection  Asymptomatic (n=1) | | First infection  Antivirals (n=1), antibiotics (n=1)  Reinfection  Antivirals (n=1) | Recovered (n=1) |
| D Harrington et al, 2020 | Case report | UK | HIC | 1 | 78 | 1 to 0 | Comorbidities (n=4) | First infection  Fever (n=1)  Reinfection  Cough (n=1), myalgia (n=1), dyspnea (n=1) | | First infection  Supportive care only  Reinfection  Steroids (n=1), antivirals (n=1), low flow oxygen (n=1) | Recovered (n=1) |
| D Larson et al, 2020 | Case report | USA | HIC | 1 | 42 | 1 to 0 | Contact history (n=1) | First infection  Fever (n=1), cough (n=1), myalgia (n=1)  Reinfection  Diarrhea (n=1), dyspnea (n=1) | | First infection  Antivirals (n=1)  Reinfection  Antivirals (n=1) | Recovered (n=1) |
| D Loconsole et al, 2020 | Case report | Italy | HIC | 1 | 48 | 1 to 0 | No contact history or comorbidities | First infection  Fever (n=1), cough (n=1), dyspnea (n=1)  Reinfection  Dyspnea (n=1) | | First infection  Antibiotics (n=1), low flow oxygen (n=1)  Reinfection  Antibiotics (n=1) | Recovered (n=1) |
| DA Torres et al, 2020 | Case report | Brazil | LMIC | 1 | 36 | 0 to 1 | No contact history or comorbidities | First infection  Fever (n=1), cough (n=1), sore throat (n=1), headache (n=1)  Reinfection  Fatigue (n=1), headache (n=1) | | First infection  Steroids (n=1), antibiotics (n=1)  Reinfection  Steroids (n=1), antibiotics (n=1) | Recovered (n=1) |
| F Bellanti et al, 2021 | Case report | Italy | HIC | 1 | 91 | 0 to 1 | Comorbidities (n=3) | First infection  Fever (n=1), cough (n=1), dyspnea (n=1)  Reinfection  Fever (n=1), dyspnea (n=1) | | First infection  Steroids (n=1), antivirals (n=1), antibiotics (n=1), low flow oxygen (n=1)  Reinfection  Steroids (n=1), antibiotics (n=1), low flow oxygen (n=1) | Deaths (n=1) |
| F Liu et al, 2020 | Case report | China | LMIC | 1 | 35 | 1 to 0 | No contact history or comorbidities | First infection  Fever (n=1), cough (n=1), fatigue (n=1), sore throat (n=1)  Reinfection  Fever (n=1), cough (n=1), sore throat (n=1) | | First infection  Steroids (n=1), antivirals (n=1), interferon (n=1)  Reinfection  Steroids (n=1), antivirals (n=1),  interferon (n=1) | Recovered (n=1) |
| FOM Alonso et al, 2021 | Case report | Brazil | LMIC | 1 | 26 | 1 to 0 | No contact history or comorbidities | First infection  Fever (n=1), myalgia (n=1), fatigue (n=1)  Reinfection  Fever (n=1), cough (n=1), myalgia (n=1), fatigue (n=1), sore throat (n=1), headache (n=1) | | First infection  Supportive care only  Reinfection  Supportive care only | Recovered (n=1) |
| G Gao et al, 2021 | Case report | China | LMIC | 1 | 70 | 1 to 0 | Comorbidities (n=1) | First infection  Fever (n=1), cough (n=1), fatigue (n=1)  Reinfection  Asymptomatic (n=1) | | First infection  Antivirals (n=1), interferon (n=1)  Reinfection  Antivirals (n=1) | Recovered (n=1) |
| G Lancman et al, 2020 | Case report | USA | HIC | 1 | 55 | 0 to 1 | Comorbidities (n=4) | First infection  Fever(n=1), cough (n=1)  Reinfection  Fever(n=1), diarrhea (n=1), sore throat (n=1) | | First infection  Antivirals (n=1), antibiotics (n=1)  Reinfection  Antibiotics (n=1),  steroids (n=1),  Low flow oxygen (n=1) | Recovered (n=1) |
| G Ye et al, 2020 | Case series | China | LMIC | 5 | 34.5 | 2 to 3 | No contact history or comorbidities | First infection  Fever (n=4), cough (n=1), fatigue (n=5)  Reinfection  Fever (n=4), cough (n=1), fatigue (n=5), sore throat (n=1) | | First infection  Steroids (n=3), antivirals (n=5), antibiotics (n=5),  Reinfection  Steroids (n=3), antivirals (n=5), antibiotics (n=5), | Recovered (n=5) |
| H Abdallah et al, 2020 | Case report | USA | HIC | 1 | 30 | 1 to 0 | Contact history (n=1) | First infection  Fever (n=1), myalgia (n=1), fatigue (n=1), dyspnea (n=1)  Reinfection  Fatigue (n=1) | | First infection  Antibiotics (n=1)  Reinfection  Antibiotics (n=1) | Recovered (n=1) |
| H Cao et al, 2020 | Case series | China | LMIC | 8 | 49 | 3 to 5 | Comorbidities (n=2) | First infection  Fever (n=5), cough (n=5), myalgia (n=1), fatigue (n=3), dyspnea (n=2)  Reinfection  Asymptomatic (n=8) | | First infection  Antivirals (n=8), traditional medicine (n=8)  Reinfection  Antivirals (n=8), traditional medicine (n=8) | Recovered (n=8) |
| H Du et al, 2020 | Case series | China | LMIC | 3 | 68 | 1 to 2 | Comorbidities (n=5) | First infection Symptoms not specified  Reinfection  Fever (n=2), cough (n=2), diarrhea (n=1), headache (n=1), dyspnea (n=1) | | First infection  Antivirals (n=3), antibiotics (n=2), traditional medicine (n=3)  Reinfection  Antivirals (n=3), antibiotics (n=2), traditional medicine (n=3) | Recovered (n=3) |
| H Zhu et al, 2020 | Case series | China | LMIC | 17 | 54 | 5 to 12 | Contact history (n=15), comorbidities (n=4) | First infection  Fever (n=12), cough (n=8), myalgia (n=7), diarrhea (n=1), fatigue (n=4)  Reinfection  Fever (n=12), cough (n=8), myalgia (n=7), diarrhea (n=1), fatigue (n=4) | | First infection  Antivirals (n=17)  Reinfection  Antivirals (n=17) | Recovered (n=17) |
| I Sicsic Jr et al, 2021 | Case report | USA | HIC | 1 | 69 | 0 to 1 | Comorbidities (n=3) | First infection  Fever (n=1), cough (n=1), fatigue (n=1), headache (n=1), dyspnea (n=1)  Reinfection  Fever (n=1), cough (n=1), dyspnea (n=1) | | First infection  Antivirals (n=1), antibiotics (n=1)  Reinfection  Steroids (n=1), antivirals (n=1), antibiotics (n=1), low flow oxygen (n=1) | Recovered (n=1) |
| J An et al, 2020 | Case series | China | LMIC | 38 | 37 | 16 to 22 | Comorbidities (n=2) | First infection  Cough (n=6), diarrhea (n=18)  Reinfection  Cough (n=6), asymptomatic (n=27) | | First infection  Supportive care only  Reinfection  Traditional medicine (n=11), interferon (n=1), low flow oxygen (n=4) | Recovered (n=38) |
| J Chen et al, 2020 | Case series | China | LMIC | 81 | 62 | 30 to 51 | Comorbidities (n=46) | First infection  Asymptomatic (n=15), rest of symptoms not specified  Reinfection  Fever (n=41), cough (n=44), myalgia (n=15), fatigue (n=34), dyspnea (n=18), asymptomatic not specified | | First infection  Antivirals (n=69)  Reinfection  Antivirals (n=69), traditional medicine (n=51), interferon (n=17), low flow oxygen (n=37) | Recovered (n=81) |
| J Goldman et al, 2020 | Case report | USA | HIC | 1 | 64.5 | 1 to 0 | Contact History (n=1), Comorbidities (n=2) | First infection,  Fever (n=1), cough (n=1), myalgia (n=1), dyspnea (n=1)  Reinfection  Cough(n=1), fatigue (n=1), dyspnea (n=1) | | First infection  Steroids (n=1), interferon (n=1), low flow oxygen (n=1)  Reinfection  Steroids (n=1), antiviral therapy (n=1) | Recovered (n=1) |
| J Huang et al, 2020 | Case series | China | LMIC | 69 | 43 | 28 to 41 | Comorbidities (n=6) | First infection symptoms not specified  Reinfection  Fever (n=43), cough (n=28), myalgia (n=2), diarrhea (n=3), fatigue (n=8), headache (n=7), dyspnea (n=1) | | First infection  Supportive care only  Reinfection  Supportive care only | Recovered (n=68), under treatment at the time of reporting (n=1) |
| J Li et al, 2020 | Case series | China | LMIC | 19 | 48 | 12 to 7 | No contact history or comorbidities | First infection  Cough (n=16)  Reinfection  Cough (n=1), dyspnea (n=1), asymptomatic (n=17) | | First infection  Steroids (n=18)  Reinfection  Steroids (n=4) | Recovered (n=19) |
| J Parry, 2020 | Case report | Hong Kong | HIC | 1 | 33 | 1 to 0 | Contact history (n=1) | First infection Symptoms not specified  Reinfection  Asymptomatic (n=1) | | First infection  Steroids (n=1), antivirals (n=1), antibiotics (n=1)  Reinfection  Steroids (n=1), antivirals (n=1), antibiotics (n=1) | Recovered (n=1) |
| J Van Elslande et al, 2020 | Case report | Belgium | HIC | 1 | 51 | 0 to 1 | Comorbidities (n=1) | First infection  Fever (n=1), cough (n=1), myalgia (n=1), headache (n=1), dyspnea (n=1),  Reinfection  Cough (n=1), headache (n=1), fatigue (n=1) | | First infection  Supportive care only  Reinfection  Supportive care only | Recovered (n=1) |
| J West et al, 2020 | Case report | UK | HIC | 1 | 25 | 1 to 0 | Contact history (n=1), no comorbidities | First infection  Fever (n=1), cough (n=1), fatigue (n=1), headache (n=1)  Reinfection  Cough (n=1), fatigue (n=1), sore throat (n=1) | | First infection  Supportive care only  Reinfection  Supportive care only | Recovered (n=1) |
| J Wu et al, 2020 | Case series | China | LMIC | 2 | 27 | 1 to 1 | Contact history (n=2) | First infection  Fever (n=2)  Reinfection  Asymptomatic (n=2) | | First infection  Steroids (n=2), antivirals (n=2), traditional medicine (n=1)  Reinfection  Steroids (n=2) | Recovered (n=2) |
| J Yuan et al, 2020 | Case series | China | LMIC | 25 | 37 | 8 to 17 | Contact history (n=25) | First infection  Fever (n=17), cough (n=15)  Reinfection  Cough (n=8), asymptomatic (n=17) | | First infection  Antivirals (n=25), traditional medicine (n=25), interferon (n=25)  Reinfection  Antivirals (n=25), traditional medicine (n=25) | Recovered (n=25) |
| J Zheng et al, 2020 | Case series | China | LMIC | 27 | 44 | 12 to 15 | Comorbidities (n=8) | First infection  Fever (n=18), cough (n=14), myalgia (n=1), fatigue (n=4), asymptomatic (n=5)  Reinfection  Fever (n=1), cough (n=6), myalgia (n=2), fatigue (n=1), sore throat (n=1) headache (n=1), asymptomatic (n=17) | | First infection  Antivirals (n=4), low flow oxygen (n=9)  Reinfection  Antivirals (n=7), low flow oxygen (n=4) | Recovered (n=27) |
| JL Moore et al, 2020 | Case report | USA | HIC | 1 | 63 | 0 to 1 | Contact history (n=1), comorbidities (n=1) | First infection  Fever (n=1), cough (n=1), myalgia (n=1)  Reinfection  Asymptomatic (n=1) | | First infection  Steroids (n=1), antibiotics (n=1)  Reinfection  Steroids (n=1), antibiotics (n=1) | Recovered (n=1) |
| K Arteaga-Livias et al, 2020 | Case report | Peru | LMIC | 1 | 42 | 0 to 1 | Contact history (n=1), comorbidities (n=1) | First infection  Cough (n=1), headache (n=1), dyspnea (n=1)  Reinfection  Fever (n=1), cough (n=1) | | First infection  Steroids (n=1), antivirals (n=1), antibiotics (n=1)  Reinfection  Supportive care only | Recovered (n=1) |
| KH Song et al, 2021 | Case series | Korea | HIC | 4 | 37.3 | 1 to 3 | Contact history (n=1), comorbidities (n=1) | First infection  Fever (n=2), cough (n=2), sore throat (n=1), headache (n=1), asymptomatic (n=1)  Reinfection  Fever (n=1), cough (n=1), asymptomatic (n=2) | | First infection  Steroids (n=4), antivirals (n=1)  Reinfection  Antivirals (n=3), antibiotics (n=1) | Recovered (n=4) |
| KI Zheng et al, 2020 | Case series | China | LMIC | 3 | Not specified | Not specified | No contact history or comorbidities | First infection symptoms not specified  Reinfection symptoms not specified | | First infection  Supportive care only  Reinfection  Supportive care only | Recovered (n=3) |
| KKW To et al, 2020 | Case report | Hong Kong | HIC | 1 | 33 | 1 to 0 | No contact history or comorbidities | First infection  Fever (n=1), cough (n=1), sore throat (n=1), headache (n=1)  Reinfection  Asymptomatic (n=1) | | First infection  Antivirals (n=1)  Reinfection  Supportive Care only | Recovered (n=1) |
| L Lafaie et al, 2020 | Case series | France | HIC | 3 | 86 | 0 to 3 | Comorbidities (n=6) | First infection  Fever (n=3), cough (n=1), myalgia (n=1), dyspnea (n=3)  Reinfection  Fever (n=2), cough (n=1), dyspnea (n=3) | | First infection  Steroids (n=3), antibiotics (n=3)  Reinfection  Steroids (n=3), antibiotics (n=3) | Deaths (n=3) |
| L Lan et al, 2020 | Case series | China | LMIC | 4 | 33 | 2 to 2 | No contact history or comorbidities | First infection  Fever (n=3), cough (n=3), asymptomatic (n=1)  Reinfection  Fever (n=3), cough (n=3), asymptomatic (n=1) | | First infection  Steroids (n=4), antivirals (n=4),  Reinfection  Steroids (n=4), antivirals (n=4), | Recovered (n=4) |
| L Pan et al, 2021 | Case series | China | LMIC | 14 | 40.5 | 10 to 4 | Comorbidities (n=3) | First infection  Fever (n=11), cough (n=10), myalgia (n=3), diarrhea (n=1), fatigue (n=7), dyspnea (n=1)  Reinfection  Antivirals (n=10), antibiotics (n=1), traditional medicine (n=3), interferon (n=4) | | First infection  Steroids (n=4), antivirals (n=14), antibiotics (n=5), traditional medicine (n=11), interferon (n=8)  Reinfection  Cough (n=1), asymptomatic (n=13) | Recovered (n=14) |
| LA Dos Santos et al, 2021 | Case series | Brazil | LMIC | 33 | 39.2 | 7 to 26 | Comorbidities (n=3) | First infection  Fever (n=7), cough (n=15), myalgia (n=16), diarrhea (n=16), sore throat (n=10), headache (n=29), dyspnea (n=10)  Reinfection  Fever (n=12), cough (n=21), myalgia (n=24), diarrhea (n=16), sore throat (n=14), headache (n=28), dyspnea (n=17) | | First infection  Steroids (n=12),  Reinfection  Steroids (n=4), low flow oxygen (n=3) | Deaths (n=1), Recovered (n=32) |
| LP Bonifácio et al, 2020 | Case report | Brazil | LMIC | 1 | 24 | 0 to 1 | Contact history (n=1)  Comorbidities (n=2) | First infection  Fever (n=1), cough (n=1), sore throat (n=1), headache (n=1)  Reinfection Fever (n=1), cough (n=1), diarrhea (n=1), sore throat (n=1), headache (n=1) | | First infection  Antibiotics (n=1)  Reinfection  Antivirals (n=1) | Recovered (n=1) |
| M Bellesso et al, 2021 | Case report | Brazil | LMIC | 1 | 76 | 0 to 1 | Comorbidities (n=2) | First infection  Fever (n=1), myalgia (n=1), fatigue (n=1)  Reinfection  Fever (n=1), myalgia (n=1), dyspnea (n=1) | | First infection  Steroids (n=1), antibiotics (n=1), interferon (n=1)  Reinfection  Steroids (n=1), antibiotics (n=1), low flow oxygen (n=1) | Deaths (n=1) |
| M Bongiovann, 2020 | Case report | Italy | HIC | 1 | 48 | 0 to 1 | Contact History (n=1) | First infection  Fever(n=1), cough (n=1)  Reinfection  Asymptomatic (n=1) | | First infection  Supportive care only  Reinfection  Supportive care only | Recovered (n=1) |
| M Gousseff et al, 2020 | Case series | France | HIC | 11 | 55 | 6 to 5 | Comorbidities (n=5) | First Infection  Fever (n=9), cough (n=5), myalgia (n=2), diarrhea (n=1), fatigue (n=6), sore throat (n=1), headache (n=5), dyspnea (n=6)  Reinfection  Fever (n=9), cough (n=5), myagia (n=2), diarrhea (n=1), fatigue (n=6), sore throat (n=1), headache (n=5), dyspnea (n=6) | | First infection  Steroids (n=3), antivirals (n=1), antibiotics (n=7), low flow oxygen (n=4)  Reinfection  Steroids (n=2), antibiotics (n=5), low flow oxygen (n=3) | Recovered (n=8), Deaths (n=3) |
| M Hanif et al, 2020 | Case report | Pakistan | LMIC | 1 | 58 | 1 to 0 | Contact History (n=1) | First infection  Fever (n=1), cough (n=1), fatigue (n=1), sore throat (n=1), headache (n=1), dyspnea (n=1)  Reinfection  Fever (n=1), myalgia (n=1), headache (n=1) | | First infection  Antibiotics (n=1), low flow oxygen (n=1)  Reinfection  Antibiotics (n=1), low flow oxygen (n=1) | Recovered (n=1) |
| M Mulder et al, 2020 | Case report | Netherlands | HIC | 1 | 89 | 0 to 1 | Comorbidities (n=1) | First infection  Fever (n=1), cough (n=1), fatigue (n=1)  Reinfection  Fever(n=1), cough (n=1), dyspnea (n=1) | | First infection  Steroids (n=1)  Reinfection  Steroids (n=1) | Deaths (n=1) |
| M Tian et al, 2020 | Case series | China | LMIC | 20 | 37.2 | 12 to 8 | Contact history (n=14), comorbidities (n=3) | First infection  Asymptomatic (n=20)  Reinfection  Asymptomatic (n=20) | | First infection  Antivirals (n=20),  interferon (n=20)  Reinfection  Antivirals (n=20),  traditional medicine (n=17), interferon (n=20) | Recovered (n=20) |
| N Zucman et al, 2021 | Case Report | France | HIC | 1 | 58 | 1 to 0 | No contact history or comorbidities | First infection  Fever (n=1), cough (n=1), dyspnea (n=1)  Reinfection  Fever (n=1), dyspnea (n=1) | | First infection  Steroids (n=1)  Reinfection  Steroids (n=1) | Recovered (n=1) |
| NM Duggan et al, 2020 | Case report | USA | HIC | 1 | 82 | 1 to 0 | Comorbidities (n=4) | First infection  Fever(n=1), myalgia (n=1)  Reinfection  Fever(n=1), dyspnea (n=1) | | First infection  Antibiotics (n=1), low flow oxygen (n=1)  Reinfection  Antibiotics (n=1),  low flow oxygen (n=1) | Recovered (n=1) |
| P Colson et al, 2020 | Case report | France | HIC | 1 | 70 | 1 to 0 | Comorbidities (n=1) | First infection  Fever (n=1), cough (n=1)  Reinfection  Asymptomatic (n=1) | | First infection  Supportive care only  Reinfection  Supportive care only | Recovered (n=1) |
| P Habibzadeh et al, 2020 | Case series | Iran | LMIC | 9 | 52 | 5 to 4 | Comorbidities (n=3) | First infection  Fever (n=9), cough (n=9), dyspnea (n=9)  Reinfection  Asymptomatic (n=9) | | First infection  Antivirals (n=9),  Reinfection  Antivirals (n=9) | Recovered (n=9) |
| P Selhorst et al, 2020 | Case report | Belgium | HIC | 1 | 27.5 | 0 to 1 | Contact history (n=1) | First infection symptoms not  specified  Reinfection  Symptoms not specified | | First infection  Antivirals (n=1)  Reinfection  Antivirals (n=1) | Recovered (n=1) |
| P Vetter et al, 2021 | Case report | Switzerland | HIC | 1 | 36 | 0 to 1 | No contact history or comorbidities | First infection  Cough (n=1), headache (n=1), dyspnea (n=1)  Reinfection  Fever (n=1), headache (n=1), dyspnea (n=1) | | First infection  Supportive care only  Reinfection  Supportive care only | Recovered (n=1) |
| PKS Chan et al, 2020 | Case report | Hong Kong | HIC | 1 | 33 | 1 to 0 | No contact history or comorbidities | First infection  Fever (n=1), cough (n=1), sore throat (n=1), headache (n=1)  Reinfection  Asymptomatic (n=1) | | First infection  Supportive care only  Reinfection  Supportive care only | Recovered (n=1) |
| Q Mei et al, 2020 | Case series | China | LMIC | 23 | 56 | 11 to 12 | Comorbidities (n=3) | First infection  Fever (n=12), cough (n=11), myalgia (n=5), fatigue (n=3), dyspnea (n=5)  Reinfection  Fever (n=6), cough (n=2), fatigue (n=1), dyspnea (n=1), asymptomatic (n=15) | | First infection  Antivirals (n=23), traditional medicine (n=23)  Reinfection  Antivirals (n=23), traditional medicine (n=23) | Recovered (n=23) |
| R Ak et al, 2021 | Case Report | Turkey | LMIC | 1 | 40 | 1 to 0 | No contact history or comorbidities | First infection  Fever (n=1), cough (n=1)  Reinfection  Cough (n=1), diarrhea (n=1), sore throat (n=1) | | First infection  Antibiotics (n=1)  Reinfection  Antibiotics (n=1) | Recovered (n=1) |
| R Kapoor et al, 2021 | Case series | India | LMIC | 3 | 32.5 | 3 to 0 | Comorbidities (n=3) | First infection  Fever (n=1), cough (n=1), asymptomatic (n=2)  Reinfection  Fever (n=3), headache (n=1), dyspnea (n=1) | | First infection  Steroids (n=2), antivirals (n=1), antibiotics (n=1)  Reinfection  Steroids (n=1), antivirals (n=1), antibiotics (n=1), low flow oxygen (n=1) | Recovered (n=3) |
| R Sharma et al, 2020 | Case report | Qatar | HIC | 1 | 57 | 1 to 0 | Contact history (n=1), comorbidities (n=1) | First infection  Asymptomatic (n=1)  Reinfection  Fever (n=1), cough (n=1), myalgia (n=1), headache (n=1) | | First infection  Antivirals (n=1)  Reinfection  Antivirals (n=1), antibiotics (n=1) | Recovered (n=1) |
| RL Tillett et al, 2020 | Case report | USA | HIC | 1 | 25 | 1 to 0 | No contact history or comorbidities | First infection  Fever (n=1), cough (n=1), myalgia (n=1), diarrhea (n=1), sore throat (n=1), headache (n=1)  Reinfection  Cough (n=1), myalgia (n=1), dyspnea (n=1) | | First infection  Steroids (n=1), antivirals (n=1)  Reinfection  Steroids (n=1), antivirals (n=1) | Recovered (n=1) |
| RP de Jesus et al, 2020 | Case report | Portugal | HIC | 1 | 41 | 1 to 0 | Contact history (n=1) | First infection  Fever (n=1), cough (n=1), myalgia (n=1), headache (n=1)  Reinfection  Fever (n=1), cough (n=1), myalgia (n=1), headache (n=1), dyspnea (n=1) | | First infection  Steroids (n=1), antivirals (n=1), antibiotics (n=1)  Reinfection  Steroids (n=1), antivirals (n=1), antibiotics (n=1) | Under treatment at the time of reporting (n=1) |
| S Atici et al, 2021 | Case series | Turkey | LMIC | 2 | 46.5 | 1 to 1 | No contact history or comorbidities | First infection  Fever (n=1), cough (n=1), myalgia (n=2), diarrhea (n=1), sore throat (n=1), headache (n=2)  Reinfection  Fever (n=2), cough (n=1), myalgia (n=1), sore throat (n=1), headache (n=1), dyspnea (n=1) | | First infection  Antibiotics (n=2), Low flow oxygen (n=1)  Reinfection  Antivirals (n=1), antibiotics (n=1) | Recovered (n=2) |
| S He et al, 2020 | Case series | China | LMIC | 30 | 66 | 17 to 13 | Comorbidities (n=7) | First infection  Fever (n=25), cough (n=25), myalgia (n=4), diarrhea (n=1), fatigue (n=6), headache (n=2)  Reinfection  Cough (n=6), fatigue (n=13), dyspnea (n=10) | | First infection  Steroids (n=18), antivirals (n=8), antibiotics (n=22), low flow oxygen (n=4)  Reinfection  Steroids (n=18), antivirals (n=8), antibiotics (n=22), low flow oxygen (n=4) | Recovered (n=30) |
| S Salcin et al, 2020 | Case report | USA | HIC | 1 | 62 | 0 to 1 | Contact history (n=1), Comorbidities (n=4) | First infection  Cough (n=1), dyspnea (n=1)  Reinfection  Asymptomatic (n=1) | | First infection  Antivirals (n=1), antibiotics (n=1)  Reinfection  Steroids (n=1), antivirals (n=1), antibiotics (n=1), low flow oxygen (n=1) | Recovered (n=1) |
| S Tomassini et al, 2020 | Case series | UK | HIC | 6 | 75.83 | 3 to 3 | Comorbidities (n=7) | First infection  Fever (n=3), cough (n=3), myalgia (n=1), diarrhea (n=2), sore throat (n=1), dyspnea (n=3)  Reinfection  Fever (n=3), cough (n=2), dyspnea (n=2), asymptomatic (n=3) | | First infection  Steroids (n=1), antibiotics (n=5), low flow oxygen (n=1)  Reinfection  Steroids (n=1), antibiotics (n=5), low flow oxygen (n=1) | Recovered (n=6) |
| S Yadav et al, 2020 | Case series | India | LMIC | 2 | 8.5 | 2 to 0 | No contact history or comorbidities | First infection  Asymptomatic (n=2)  Reinfection  Asymptomatic (n=2) | | First infection  Supportive care only (n=2)  Reinfection  Supportive care only (n=2) | Recovered (n=2) |
| S Zayet et al, 2021 | Case series | France | HIC | 3 | 44.5 | 0 to 3 | Comorbidities (n=1) | First infection  Fever (n=2), cough (n=1), myalgia (n=3), fatigue (n=3), sore throat (n=3)  Reinfection  Fever (n=2), cough (n=2), myalgia (n=2), diarrhea (n=1), fatigue (n=2), headache (n=2), dyspnea (n=2) | | First infection  Supportive care only  Reinfection  Supportive care only | Recovered (n=3) |
| SP Yadav et al, 2021 | Case report | India | LMIC | 1 | 3 | 1 to 0 | Comorbidities (n=1) | First infection  Asymptomatic (n=1),  Reinfection  Asymptomatic (n=1) | | First infection  Supportive care only  Reinfection  Supportive care only | Recovered (n=1) |
| SY Yoo et al, 2020 | Case report | China | LMIC | 1 | 8 | 1 to 0 | Contact history (n=1) | First infection  Cough (n=1)  Reinfection  Fever (n=1) | | First infection  Supportive care only  Reinfection  Supportive care only | Recovered (n=1) |
| V Gupta et al, 2020 | Case series | India | LMIC | 2 | 26.5 | 1 to 1 | Contact history (n=2) | First infection  Asymptomatic (n=2)  Reinfection  Asymptomatic (n=2) | | First infection  Supportive care only  Reinfection  Supportive care only | Recovered (n=2) |
| V Nachmias et al, 2020 | Case report | Israel | HIC | 1 | 20 | 0 to 1 | Contact history (n=1) | First infection  Fever (n=1), cough (n=1)  Reinfection  Asymptomatic (n=1) | | First infection  Antiviral therapy (n=1)  Reinfection  Supportive care only | Recovered (n=1) |
| V Selvaraj et al, 2020 | Case report | USA | HIC | 1 | 70 | 1 to 0 | Contact history (n=1), comorbidities (n=6) | First infection  Fever (n=1), dyspnea (n=1)  Reinfection  Fever (n=1), myalgia (n=1), dyspnea (n=1) | | First infection  Antivirals (n=1), antibiotics (n=1)  Reinfection  Steroids (n=1), antivirals (n=1), antibiotics (n=1), low flow oxygen (n=1) | Recovered (n=1) |
| W Fu et al, 2020 | Case series | China | LMIC | 3 | 48 | 1 to 2 | Comorbidities (n=1) | First infection  Fever (n=3), cough (n=2)  Reinfection  Asymptomatic (n=3) | | First Infection  Antivirals (n=3), antibiotics (n=3), traditional medicine (n=3)  Reinfection  Antivirals (n=3), antibiotics (n=3), | Recovered (n=3) |
| W Novoa et al, 2021 | Case report | Colombia | LMIC | 1 | 44 | 1 to 0 | No contact history or comorbidities | First infection  Asymptomatic (n=1)  Reinfection  Fever (n=1), cough (n=1), myalgia (n=1), fatigue (n=1), sore throat (n=1), headache (n=1) | | First infection  Supportive care only  Reinfection  Antibiotics (n=1) | Recovered (n=1) |
| W Zhao et al, 2020 | Case series | China | LMIC | 7 | 8.5 | 3 to 4 | No contact history or comorbidities | First infection  Fever (n=5), cough (n=1), headache (n=1), asymptomatic (n=2)  Reinfection  Fever (n=3), cough (n=1), diarrhea (n=1), asymptomatic (n=2) | | First infection  Antivirals (n=3), traditional medicine (n=4)  Reinfection  Antivirals (n=4), traditional medicine (n=3) | Recovered (n=7) |
| X Li et al, 2020 | Case report | China | LMIC | 1 | 41 | 1 to 0 | No contact history or comorbidities | First infection  Fever (n=1)  Reinfection  Cough (n=1) | | First infection  Antiviral therapy (n=1), traditional medicine (n=1), interferon (n=1), low flow oxygen (n=1)  Reinfection  Traditional medicine (n=1), low flow oxygen (n=1) | Recovered (n=1) |
| Y Li et al, 2020 | Case series | China | LMIC | 4 | 55 | 2 to 2 | Comorbidities (n=4) | First infection  Fever (n=4), cough (n=4), myalgia (n=1), fatigue (n=2)  Reinfection  Asymptomatic (n=4) | | First infection  Antivirals (n=4), antibiotics (n=2), low flow oxygen (n=3)  Reinfection  Antivirals (n=4), antibiotics (n=2), low flow oxygen (n=3) | Recovered (n=4) |
| Y Ling et al, 2020 | Case series | China | LMIC | 11 | 44 | 5 to 6 | No contact history or comorbidities | First infection symptoms not specified  Reinfection  Fever (n=11), cough (n=11), dyspnea (n=11) | | First infection  Steroids (n=5)  Reinfection  Steroids (n=5) | Recovered (n=11) |

NR= Not reported
