## Supplementary material for "The mystery of COVID-19 reinfections: A global systematic review and meta-analysis of 577 cases": S3 Table. Quality assessment of case reports and case series (N=81)

| **Study and Year** | **1. Was the study question or objective clearly stated?** | **2. Was the study population clearly and fully described, including a case definition?** | **3. Were the cases consecutive?** | **4. Were the subjects comparable?** | **5. Was the intervention clearly described?** | **6. Were the outcome measures clearly defined, valid, reliable, and implemented consistently across all study participants?** | **7. Was the length of follow-up adequate?** | **8. Were the statistical methods well-described?** | **9. Were the results well-described?** | **Quality Rating (Good, Fair, or Poor)** |
| --- | --- | --- | --- | --- | --- | --- | --- | --- | --- | --- |
| A Coppola et al, 2020 | Yes | Yes | NA | NA | NA | Yes | Yes | NA | Yes | Good |
| A Luo, 2020 | Yes | Yes | NA | NA | NA | Yes | Yes | NA | Yes | Good |
| AM Ali et al, 2020 | Yes | Yes | Yes | Yes | NA | Yes | Yes | Yes | Yes | Good |
| B Prado-Vivar et al, 2020 | Yes | Yes | NA | NA | NA | Yes | Yes | NA | Yes | Good |
| B Zhang et al, 2020 | Yes | Yes | No | Yes | NA | Yes | Yes | Yes | Yes | Good |
| C Dou et al, 2020 | Yes | Yes | NA | NA | NA | Yes | Yes | NA | Yes | Good |
| CAA de Brito et al, 2020 | Yes | Yes | No | Yes | NA | Yes | Yes | Yes | Yes | Good |
| CFV Takeda et al, 2020 | Yes | Yes | No | Yes | NA | Yes | Yes | Yes | Yes | Good |
| D Chen et al, 2020 | Yes | Yes | NA | NA | NA | Yes | Yes | NA | Yes | Good |
| D Harrington et al, 2020 | Yes | Yes | NA | NA | NA | Yes | Yes | NA | Yes | Good |
| D Larson et al, 2020 | Yes | No | NA | NA | NA | Yes | Yes | NA | Yes | Fair |
| D Loconsole et al, 2020 | Yes | Yes | NA | NA | NA | Yes | Yes | NA | Yes | Good |
| DA Torres et al, 2020 | Yes | Yes | NA | NA | NA | Yes | Yes | NA | Yes | Good |
| F Bellanti et al, 2021 | Yes | Yes | NA | NA | NA | Yes | Yes | NA | Yes | Good |
| F Liu et al, 2020 | Yes | Yes | NA | NA | NA | Yes | Yes | NA | Yes | Good |
| FOM Alonso et al, 2021 | Yes | No | NA | NA | NA | Yes | Yes | NA | Yes | Fair |
| G Gao et al, 2021 | Yes | Yes | NA | NA | NA | Yes | Yes | NA | Yes | Good |
| G Lancman et al, 2020 | Yes | Yes | NA | NA | NA | Yes | Yes | NA | Yes | Good |
| G Ye et al, 2020 | Yes | Yes | Yes | Yes | NA | Yes | Yes | Yes | Yes | Good |
| H Abdallah et al, 2020 | Yes | Yes | NA | NA | NA | Yes | Yes | NA | Yes | Good |
| H Cao et al, 2020 | Yes | Yes | Yes | Yes | NA | Yes | Yes | Yes | Yes | Good |
| H Du et al, 2020 | Yes | Yes | Yes | Yes | NA | Yes | Yes | Yes | Yes | Good |
| H Zhu et al, 2020 | Yes | Yes | No | Yes | NA | Yes | Yes | Yes | Yes | Good |
| I Sicsic Jr et al, 2021 | Yes | Yes | NA | NA | NA | Yes | Yes | NA | Yes | Good |
| J An et al, 2020 | Yes | Yes | Yes | Yes | NA | Yes | Yes | Yes | Yes | Good |
| J Chen et al, 2020 | Yes | Yes | No | Yes | NA | Yes | Yes | Yes | Yes | Good |
| J Goldman et al, 2020 | Yes | Yes | NA | NA | NA | Yes | Yes | NA | Yes | Good |
| J Huang et al, 2020 | Yes | Yes | Yes | Yes | NA | Yes | No | Yes | Yes | Good |
| J Li et al, 2020 | Yes | Yes | No | Yes | NA | Yes | Yes | Yes | Yes | Good |
| J Parry, 2020 | Yes | Yes | NA | NA | NA | Yes | Yes | NA | Yes | Good |
| J Van Elslande et al, 2020 | Yes | Yes | NA | NA | NA | Yes | Yes | NA | Yes | Good |
| J West et al, 2020 | Yes | Yes | NA | NA | NA | Yes | Yes | NA | No | Fair |
| J Wu et al, 2020 | Yes | Yes | No | No | NA | Yes | Yes | Yes | Yes | Good |
| J Yuan et al, 2020 | Yes | Yes | Yes | Yes | NA | Yes | Yes | Yes | Yes | Good |
| J Zheng et al, 2020 | Yes | Yes | Yes | Yes | NA | Yes | Yes | Yes | Yes | Good |
| JL Moore et al, 2020 | Yes | Yes | NA | NA | NA | Yes | Yes | NA | Yes | Good |
| K Arteaga-Livias et al, 2020 | Yes | Yes | NA | NA | NA | Yes | Yes | NA | Yes | Good |
| KH Song et al, 2021 | Yes | No | Yes | Yes | NA | Yes | Yes | Yes | Yes | Good |
| KI Zheng et al, 2020 | Yes | Yes | Yes | Yes | NA | Yes | Yes | Yes | Yes | Good |
| KKW To et al, 2020 | Yes | Yes | NA | NA | NA | Yes | Yes | NA | Yes | Good |
| L Lafaie et al, 2020 | Yes | Yes | No | Yes | NA | Yes | Yes | Yes | Yes | Good |
| L Lan et al, 2020 | Yes | Yes | No | Yes | NA | Yes | Yes | No | Yes | Good |
| L Pan et al, 2021 | Yes | Yes | Yes | Yes | NA | Yes | Yes | Yes | Yes | Good |
| LA Dos Santos et al, 2021 | Yes | Yes | Yes | Yes | NA | Yes | Yes | Yes | Yes | Good |
| LP Bonifácio et al, 2020 | Yes | Yes | NA | NA | NA | Yes | Yes | NA | Yes | Good |
| M Bellesso et al, 2021 | Yes | Yes | NA | NA | NA | Yes | Yes | NA | Yes | Good |
| M Bongiovanni, 2020 | Yes | Yes | NA | NA | NA | Yes | Yes | NA | Yes | Good |
| M Gousseff et al, 2020 | Yes | Yes | Yes | Yes | NA | Yes | Yes | Yes | Yes | Good |
| M Hanif et al, 2020 | Yes | Yes | NA | NA | NA | Yes | Yes | NA | Yes | Good |
| M Mulder et al, 2020 | Yes | Yes | NA | NA | NA | Yes | Yes | NA | Yes | Good |
| M Tian et al, 2020 | Yes | Yes | Yes | Yes | NA | Yes | Yes | Yes | Yes | Good |
| N Zucman et al, 2021 | Yes | Yes | NA | NA | NA | Yes | Yes | NA | Yes | Good |
| NM Duggan et al, 2020 | Yes | Yes | NA | NA | NA | Yes | Yes | NA | Yes | Good |
| P Colson et al, 2020 | Yes | No | NA | NA | NA | Yes | Yes | NA | Yes | Fair |
| P Habibzadeh et al, 2020 | Yes | Yes | No | Yes | NA | Yes | Yes | No | No | Fair |
| P Selhorst et al, 2020 | Yes | No | Yes | Yes | NA | Yes | Yes | NA | Yes | Good |
| P Vetter et al, 2021 | Yes | No | NA | NA | NA | Yes | Yes | NA | Yes | Fair |
| PKS Chan et al, 2020 | Yes | Yes | NA | NA | NA | Yes | Yes | NA | Yes | Good |
| Q Mei et al, 2020 | Yes | Yes | Yes | Yes | NA | Yes | Yes | Yes | Yes | Good |
| R Ak et al, 2021 | Yes | Yes | NA | NA | NA | Yes | Yes | NA | Yes | Good |
| R Kapoor et al, 2021 | Yes | Yes | No | Yes | NA | Yes | Yes | Yes | Yes | Good |
| R Sharma et al, 2020 | Yes | Yes | NA | NA | NA | Yes | Yes | NA | Yes | Good |
| RL Tillett et al, 2020 | Yes | Yes | NA | NA | NA | Yes | Yes | NA | Yes | Good |
| RP de Jesus et al, 2020 | No | Yes | NA | NA | NA | Yes | No | NA | Yes | Fair |
| S Atici et al, 2021 | Yes | Yes | No | Yes | NA | Yes | Yes | Yes | Yes | Good |
| S He et al, 2020 | Yes | Yes | Yes | Yes | NA | Yes | Yes | Yes | Yes | Good |
| S Salcin et al, 2020 | Yes | Yes | NA | NA | NA | Yes | Yes | NA | Yes | Good |
| S Tomassini et al, 2020 | Yes | Yes | Yes | Yes | NA | Yes | Yes | Yes | Yes | Good |
| S Yadav et al, 2020 | Yes | Yes | No | Yes | NA | Yes | Yes | No | Yes | Good |
| S Zayet et al, 2021 | Yes | Yes | Yes | Yes | NA | Yes | Yes | Yes | Yes | Good |
| SP Yadav et al, 2021 | Yes | Yes | NA | NA | NA | Yes | Yes | NA | Yes | Good |
| SY Yoo et al, 2020 | Yes | No | NA | NA | NA | Yes | Yes | NA | Yes | Fair |
| V Gupta et al, 2020 | Yes | Yes | No | Yes | NA | Yes | Yes | NA | Yes | Good |
| V Nachmias et al, 2020 | Yes | Yes | NA | NA | NA | Yes | Yes | NA | Yes | Good |
| V Selvaraj et al, 2020 | Yes | No | NA | NA | NA | Yes | Yes | NA | Yes | Fair |
| W Fu et al, 2020 | Yes | Yes | No | Yes | NA | Yes | Yes | Yes | Yes | Good |
| W Novoa et al, 2021 | Yes | Yes | NA | NA | NA | Yes | Yes | NA | Yes | Good |
| W Zhao et al, 2020 | Yes | Yes | Yes | Yes | NA | Yes | Yes | Yes | Yes | Good |
| X Li et al, 2020 | Yes | Yes | NA | NA | NA | Yes | Yes | NA | Yes | Good |
| Y Li et al, 2020 | Yes | Yes | Yes | Yes | NA | Yes | Yes | Yes | Yes | Good |
| Y Ling et al, 2020 | Yes | Yes | Yes | Yes | NA | Yes | Yes | Yes | Yes | Good |
